# A job exposure matrix for occupational exposure to airborne micro and nanoplastics (PlastiXJEM®) and associations with respiratory outcomes

**DOI:** 10.64898/2026.03.14.26348371

**Authors:** Gwenda F. Vasse, Nienke Vrisekoop, Joëlle A.Z. Klazen, Judith M. Vonk, Barbro N. Melgert

## Abstract

**Background:** Microplastics and nanoplastics (MNP) are an increasingly recognized component of airborne particulate matter, yet their impact on respiratory health is unclear. This study aimed to develop a job exposure matrix (JEM) for occupational exposure to airborne MNP (PlastiXJEM®) and examine its association with respiratory outcomes in the Lifelines cohort.

**Methods:** Four experts scored occupational airborne MNP exposure levels (none, low, high) for all ISCO-08 occupations based on documented sources and published evidence. After consensus, the PlastiXJEM® was applied to baseline current or last-held jobs of 136,928 adult Lifelines participants. Cross-sectional and longitudinal associations with lung function, respiratory symptoms, and asthma were assessed using linear and logistic regression models adjusted for age, sex, smoking, height, BMI, and co-exposure to organic dust, gasses and fumes, pesticides, metals, solvents and silica.

**Results:** High exposure was associated with lower FEV₁ (–43 ml; 95% CI:–61;–25), lower FVC (–47 ml (–69;–26)), lower FEV1%FVC (-0.26 % (-0.51;-0.00) and higher odds of airway obstruction, respiratory symptoms and asthma (e.g. dyspnea OR=1.58; 1.34–1.87). Low exposure was associated with lower FEV_1_ and FVC in females only. Overall, effect sizes were larger at higher exposure levels, consistent with a dose-dependent pattern. MNP exposure was not associated with accelerated lung function decline or with the development of airway obstruction, respiratory symptoms, or asthma.

**Conclusion:** Occupational exposure to airborne MNP is associated with lower lung function and a higher prevalence of respiratory symptoms in this cohort. These findings warrant further investigation with complete occupational histories.

## Introduction

Plastic production and use have grown exponentially over the past decades and continue to rise at an accelerating rate [1]. Through abrasion during production and use and degradation after disposal, these plastics fragment into micro-and nanoplastics (MNP), leading to widespread environmental contamination [2, 3]. MNP are generally defined as particles and fibres with at least one external dimension between 1 µm and 5 mm and nanoplastics as particles below 1 µm, consisting of synthetic or heavily modified polymeric materials such as polyesters, polyolefins, polyamides, acrylics and synthetic rubbers [4]. This operational definition includes tyre and other synthetic rubber wear particles when they contain a predominant polymer fraction. MNP are now also recognized as an emerging component of airborne particulate matter [5–9], with the highest concentrations measured indoors, particularly in workplaces where plastic materials are processed or handled [10]. Recent studies estimate that at least 50% of human MNP intake occurs through inhalation [11, 12] and this suggests that workers in certain occupations may represent the population with the highest expected lung exposure levels. Yet, health risks from occupational inhalation remain largely unexplored.

Evidence from specific industries, such as plastic manufacturing and synthetic textile processing, signals a higher risk of respiratory health effects, including airway inflammation, lower lung function, and even lung cancer and interstitial lung diseases [2, 10, 13, 14]. Workplace studies further support that inhalation exposures can be substantially higher in occupational settings than in the general population [15, 16]. For example, airborne MNP concentrations in certain industrial processes such as 3D-printing extrusion and nylon/polypropylene flocking have been reported at levels roughly 10–100 times higher than those measured in ambient or indoor air [12, 16–25], and personal air sampling in waste-handling occupations has found thousands of inhalable MNP particles per cubic meter compared with background levels typically in the range of 1–500 particles/m³ outdoors and 1–1,200 particles/m³ indoors [12, 25, 26]. A small cross-sectional study in plastic products manufacturing workers found higher MNP counts on hair, skin and saliva after a single work shift than before, indicating direct work-related exposure [27]. Despite such indications, and the likelihood that adverse respiratory effects manifest earlier and more strongly in occupationally exposed populations, systematic epidemiological studies remain scarce, and standardized methods for assessing airborne MNP exposure are not yet available. Together, these factors highlight an urgent need for occupational exposure assessment frameworks to support large-scale epidemiological research on MNP-related respiratory health risks.

To address this, we developed a semi-quantitative, group-based job exposure matrix (JEM) for airborne MNP exposure (PlastiXJEM®) using the International Standard Classification of Occupations 2008 (ISCO-08) classification [28] and applied it to the Lifelines cohort, a large multigenerational population study in the northern Netherlands [29]. We aimed to examine cross-sectional and longitudinal associations between occupational MNP exposure and respiratory health, including lung function, airway obstruction, respiratory symptoms and asthma, with additional sex-stratified analyses given the known influence of biological sex on lung structure and susceptibility to inhaled pollutants [30, 31].

## Methods

### Development of the MNP job exposure matrix (PlastiXJEM®)

Given the lack of occupational exposure measurement data for airborne MNP, we developed a JEM based on expert judgment, a common approach in occupational epidemiology when direct measurements are unavailable. The JEM was constructed using the International Standard Classification of Occupations 2008 (ISCO-08), a globally accepted framework for classifying jobs based on duties, tasks, and required skill levels (https://ilostat.ilo.org/methods/concepts-and-definitions/classification-occupation/) [28] The list with example occupations for each job code provided by Statistics Netherlands (CBS) was used to guide consistent interpretation of each job code (Codelijsten ISCO-08 | CBS). Four independent experts in microplastic research (Barbro Melgert, Gwenda Vasse, Joëlle Klazen, and Nienke Vrisekoop) independently rated each ISCO-08 occupation for expected occupational exposure to airborne MNP. Exposure levels were categorized as follows: 0 for no occupational exposure, 1 for low occupational exposure, and 2 for high occupational exposure. Ratings were based on peer-reviewed literature regarding known sources of MNP (e.g., textile fibers, synthetic cleaning agents, 3D printing filaments, tire wear), studies on MNP concentrations in indoor and outdoor air, and the release of MNP during specific occupational activities (see Supplementary table S1 for the literature sources that informed the scoring).

Specifically, MNP were defined as all synthetic polymer particles and fibres in the micro and nano size range (e.g. polyesters, polyolefins, polyamides, acrylics and synthetic rubbers, including tyre and rubber wear when polymer based in sizes from a few nanometers to max 5 millimeters).

In cases where a job category included several sub-occupations with varying potential exposure levels, the most representative exposure level for the category as a whole was assigned based on expert consensus. In exceptional cases where a specific sub-occupation clearly deviated from the rest of its category, it was reclassified as a separate group within the JEM. Following the initial independent scoring, a structured consensus process was implemented to improve consistency and resolve discrepancies. Discrepancies were defined as any occupation where not all four raters assigned the same score, and all such occupations were discussed. During consensus meetings, each expert presented their rationale, supported by scientific literature and known exposure pathways. Scores were revised where appropriate based on these discussions.

### Study population

To investigate the association between the JEM-based occupational MNP exposure and respiratory outcomes, data from the Lifelines cohort study were used. Lifelines is a multi-disciplinary prospective population-based cohort study examining in a unique three-generation design the health and health-related behaviours of 167,729 persons living in the North of the Netherlands. It employs a broad range of investigative procedures in assessing the biomedical, socio-demographic, behavioural, physical and psychological factors which contribute to the health and disease of the general population, with a special focus on multi-morbidity and complex genetics. The Medical Ethics Committee (METc) of the University Medical Center Groningen, The Netherlands approved the study under approval number METC 2007/152. All participants provided written informed consent. [29]. Since its inception in 2006, Lifelines has collected current or last held job and health data through questionnaires, physical measurements, and biological samples during a baseline visit. Participants complete questionnaires every 1.5 years and undergo detailed health assessments every five years.

### Respiratory health outcomes

Lung function was measured by pre-bronchodilator spirometry in a random subset of the participants at baseline and at two follow-up health assessments (mean follow-up time was 8.5 years (standard deviation: 3.5). Spirometry was performed using a Welch Allyn SpiroPerfect device (Version 1.6.0.489, PC-based SpiroPerfect with CardioPerfect Workstation software; Welch Allyn) according to American Thoracic Society and European Respiratory Society guidelines [32].

Respiratory symptoms and asthma (see supplementary table S2 for the definition of the symptoms) were measured by questionnaire at baseline and at the first follow-up health assessment.

### Occupational data

At baseline, participants reported their current or last held job (in case they currently did not work) and their activities on the job in the questionnaire. All occupations were manually coded according to the International Standard Classification of Occupations version 2008 (ISCO-08) [28], using all available information, which could result in more than one code being assigned to a participant. For example, if a participant worked in a supermarket and reported both cashier and shelf-filling activities, both occupations were coded.

### Exposure assessment and assignment

The developed PlastiXJEM® was linked to the ISCO-08 codes of the current or last held job at baseline to determine occupational MNP exposure. For individuals who reported multiple occupations, the highest exposure score across all jobs was used in the analyses. For illustration purposes, occupations classified as having low or high MNP exposure were additionally grouped into broader occupational sectors (Supplementary table 2).

### Covariates

Age and sex were noted at baseline. Height and weight were measured during the baseline visit. Self-reported smoking status, education, and monthly income were extracted from the baseline questionnaires (see Supplementary table S3 for definitions). Occupational co-exposures were based on the updated version of the expert-based occupational asthma JEM (OAsJEM) [33]: organic dust exposure was defined as the highest OAsJEM-based exposure level of animals, flour, foods, plants-related dusts, moulds and endotoxin; gasses and fumes exposure was defined as the highest level of exhaust fumes, high-level chemical disinfectants and bleach; pesticides exposure was defined as the highest level of herbicides, insecticides and fungicides; metals exposure was defined as the highest level of metal and metal working fluids; and for solvents exposure the exposure of organic solvents was used. In addition, exposure to respirable crystalline silica was based on the EUROJEM [34]. For this, we used the exposure prevalence estimates for Western Europe for 2000–2009 (corresponding to the Lifelines baseline period). Following the EuroJEM instructions, we categorised silica exposure as none (group 0), low (group 1), and high (groups 2–3).

Since the OAsJEM and the EUROJEM use ISCO-88 codes, the conversion table provided by the International Labour Organization was used to convert ISCO-08 to ISCO-88 codes. Importantly, some categories were combined in ISCO-08 compared to ISCO-88 (e.g., ISCO-08 8160 - Food and related product machine operators - had different codes in ISCO-88 depending on the type of food/product - ISCO-88 8271 till 8279). For these combined categories, the jobs were manually recoded to the correct ISCO-88 category.

## Statistical analyses

The inter-rater reliability of the PlastiXJEM® scoring was assessed using Fleiss’ kappa before and after the consensus-building process. Scores were analyzed in R studio version 2024.12.0+467. Spearman rank correlations were calculated between occupational MNP exposure and other occupational exposures for the total population and stratified by sex.Descriptive statistics were calculated for the population with MNP exposure data available and also separately for the populations included in the cross-sectional longitudinal analyses. Continuous variables are presented as mean and standard deviation (SD) and categorical data are presented as frequency and percentage. Descriptives were calculated for the total population and stratified by occupational MNP exposure and by gender. Cross-sectional analyses at baseline were performed on the forced expiratory volume in 1 second (FEV1), the forced vital capacity (FVC), the FEV1 as a percentage of FVC (FEV1%FVC), airway obstruction (defined as FEV1/FVC < 0.7 and as FEV1/FVC < lower limit of normal according to the Global Lung Function Initiative reference equations [35]), wheeze, cough, phlegm, bronchitis, dyspnea, and asthma. For the continuous outcomes, linear regression was used and for the categorical outcomes logistic regression was used. Longitudinal analyses using linear mixed effect models with a random effect on the subject-specific intercept and including interactions between MNP exposure at baseline and time since baseline were performed to investigate the association between the occupational MNP exposure at baseline and the subsequent annual decline in FEV1, FVC and FEV1%FVC. In this analysis, only subjects aged 25 and older at baseline were included since the natural age-related decline in lung function starts around age 25 and only subjects with at least one follow-up measurement were included.

To investigate the association between occupational MNP exposure at baseline and the development of airway obstruction, respiratory symptoms or asthma, only subjects without airway obstruction, respiratory symptoms, or asthma at baseline were included in a logistic regression analysis on airway obstruction, respiratory symptoms, or asthma at follow-up.

The base model was adjusted for age, sex, smoking, and height (outcome: lung function) or BMI (outcome: symptoms/asthma). The main model was additionally adjusted for co-exposures to organic dust, gasses and fumes, pesticides, metals, solvents and silica. In a sensitivity analysis, additional adjustment for education and income was performed to investigate if the associations are independent of socio-economic status. In the mixed effect models on lung function decline, interactions between all covariates and time since baseline were included. The models on development of airway obstruction were additionally adjusted for time since baseline. Finally, all analyses were performed for the total cohort and separately for females and males.

## Results

### Reliability and consistency of the PlastiXJEM®

The initial scoring indicated fair agreement among the raters with a Fleiss’ kappa of 0.39 (Figure 1). Raters 2 and 4 showed the biggest disagreement, while raters 1 and 3 agreed most (supplementary Figure S1). However, most disagreements concerned only one deviating score out of four raters (in around 300 out of 436 total ISCO-08 job-codes (70%)), while two or three disagreeing scores were far less common (respectively around 110 cases (25%) and around 20 cases (5%), supplemental data Figure S2). Jobs such as’Metal moulders and coremakers’,’Painters and related workers’, and‘Beauticians and related workers’ had the greatest initial discrepancies. Based on evidence that nail styling generates microplastic dust from acrylic nails [20], nail stylists were separated from the broader’Beauticians and related workers’ category and classified as a distinct group within the JEM. After the consensus process, full agreement was reached for all occupations (Fleiss’ kappa = 1.0).

**Figure 1:**
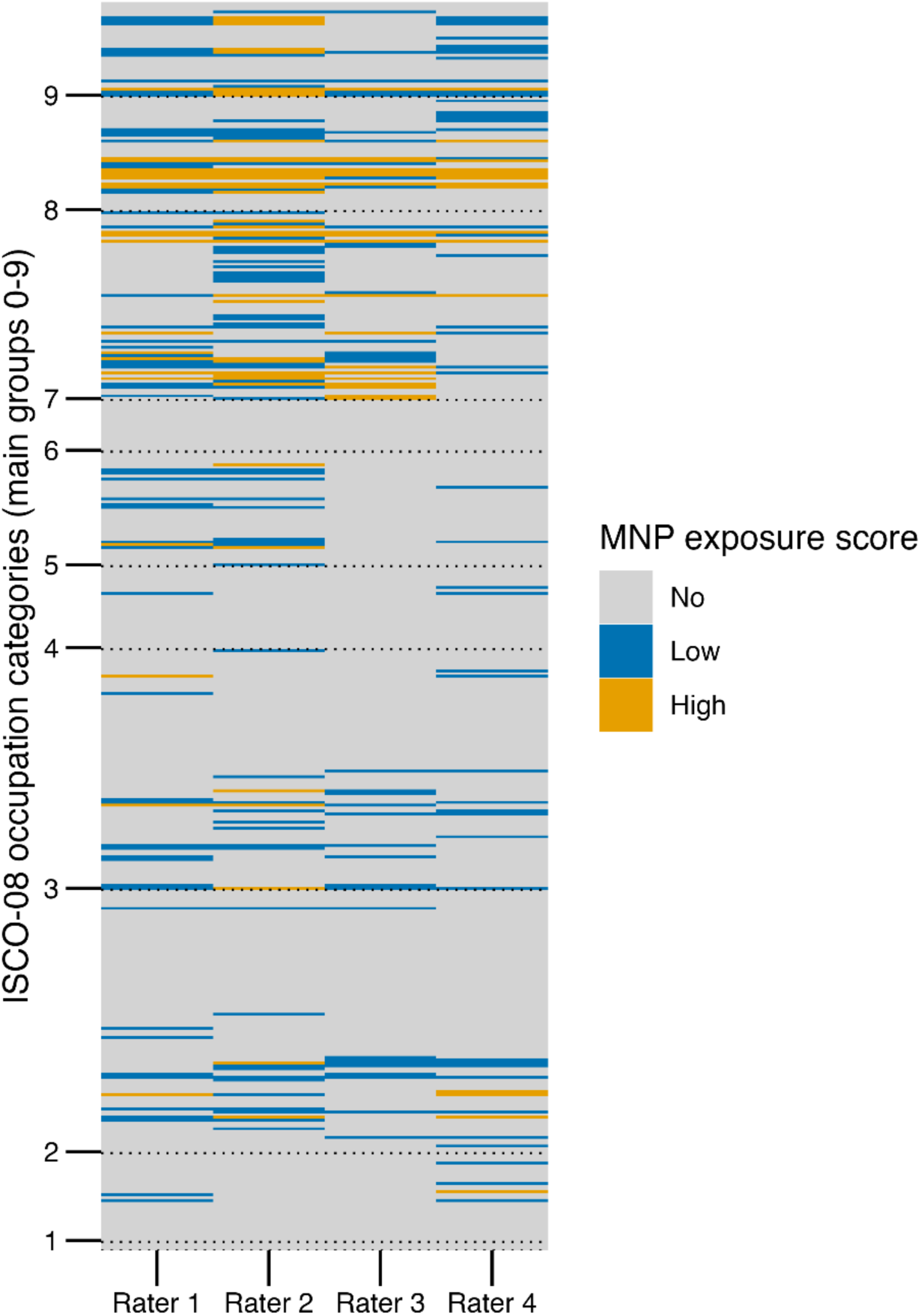
Heatmap of initial rater scores for anticipated occupational microplastic exposure. Each tile shows the score assigned by an individual rater for a given ISCO-08 occupation code. Colours denote score outcome: light gray=no, blue=low, orange=high MNP exposure.

### Study population

Having established the PlastiXJEM®, we linked it to the Lifelines cohort. The flow chart in Figure 2 shows the number of participants included in each analysis with available occupational information and relevant health data.

**Figure 2:**
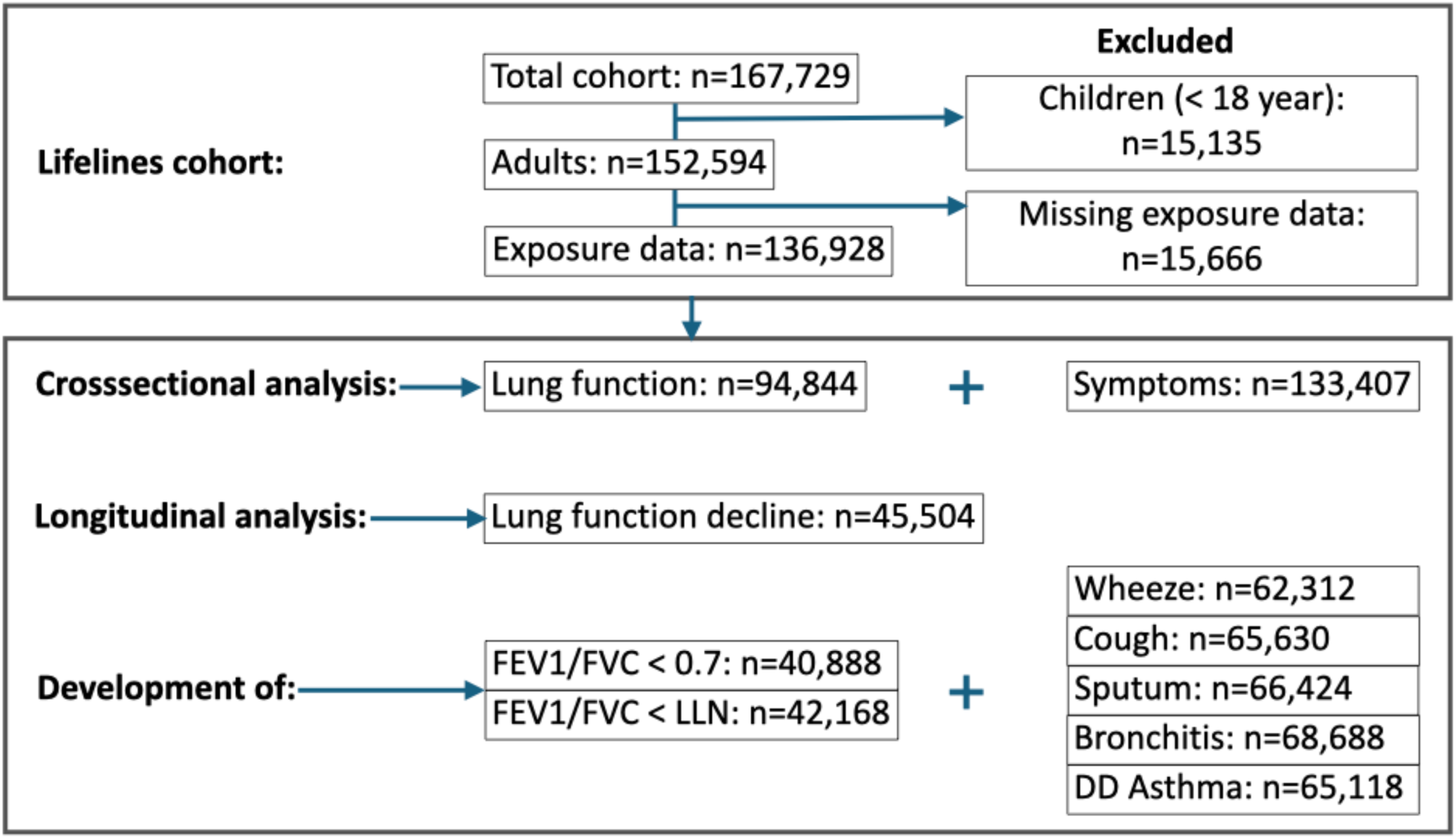
Flow chart of participant inclusion for each respiratory health analysis. The flow chart shows the number of Lifelines participants included in analyses on lung function, respiratory symptoms, asthma, and longitudinal change, based on available data and inclusion criteria.

Descriptive statistics of the subjects with exposure data available (n=136,928) are presented in Table 1. Among these, 75.1%, 21.7% and 3.2% were classified as having respectively no, low and high occupational exposure to airborne MNP. Descriptive statistics for participants included in the analyses of lung function, respiratory symptoms and longitudinal outcomes are provided in supplementary tables S4-S7.

**Table 1.** Descriptives statistics stratified by MNP exposure for participating subjects with available exposure data. Mean (SD) for continuous variables, N (%) for categorical variables. BMI: body mass index.

|  | All | No exposure | Low exposure | High exposure |
| --- | --- | --- | --- | --- |
| **N** | 136928 | 102857 (75.1) | 29659 (21.7) | 4412 (3.2) |
| **Age, year** | 44.7 (13.3) | 44.4 (13.3) | 45.5 (13.2) | 45.8 (13.1) |
| **Male sex** | 53256 (38.9) | 39872 (38.8) | 11237 (37.9) | 2147 (48.7) |
| **Height, cm** | 174.4 (9.4) | 174.6 (9.3) | 173.7 (9.3) | 174.3 (9.8) |
| **Weight, kg** | 79.3 (15.3) | 79.1 (15.1) | 79.7 (15.6) | 81.4 (16.1) |
| **BMI, kg/m<sup>2</sup>** | 26.0 (4.4) | 25.9 (4.3) | 26.4 (4.5) | 26.7 (4.7) |
| **Smoking** |  |  |  |  |
| **Never** | 60699 (44.3) | 46658 (45.4) | 12316 (41.5) | 1725 (39.1) |
| **Ex** | 43226 (31.6) | 32180 (31.3) | 9706 (32.7) | 1340 (30.4) |
| **Current** | 26918 (19.7) | 19371 (18.8) | 6412 (21.6) | 1135 (25.7) |
| **Missing** | 6085 (4.4) | 4648 (4.5) | 1225 (4.1) | 212 (4.8) |
| **Education** |  |  |  |  |
| **Low** | 41933 (30.7) | 26168 (25.4) | 13156 (44.4) | 2669 (60.5) |
| **Medium** | 53288 (38.9) | 41242 (40.1) | 10735 (36.2) | 1311 (29.7) |
| **High** | 39081 (28.5) | 33590 (32.7) | 5149 (17.4) | 342 (7.8) |
| **Missing** | 2566 (1.9) | 1857 (1.8) | 619 (2.0) | 90 (2.0) |
| **Income** |  |  |  |  |
| **Low** | 21267 (15.5) | 15367 (14.9) | 5020 (16.9) | 880 (19.9) |
| **Medium** | 34849 (25.5) | 24580 (23.9) | 9021 (30.4) | 1248 (28.3) |
| **High** | 56173 (41.0) | 45434 (44.2) | 9601 (32.4) | 1138 (25.8) |
| Unknown | 24639 (18.0) | 17476 (17.0) | 6017 (20.3) | 1146 (26.0) |

Occupational sectors with potential exposure to airborne MNP vary considerably in both their prevalence and sex distribution. Among the subjects with exposure data available 34,071 had low or high occupational exposure to MNP and of these people the largest group worked in the medical and dental sector (25.9% of all exposed people), followed by cleaning (23.8% of all exposed people) and construction (17.3% of all exposed people). This overall pattern, however, masks substantial differences between males and females. Females with exposure were predominantly employed in healthcare-related (37.6% of all exposed females) or cleaning occupations (36.7% of all exposed females), whereas males were most commonly found in construction-related jobs (43.6% of all exposed men). These sector-specific patterns illustrate how MNP exposure at work is not only shaped by occupation, but also by gendered labor participation (Figure 3).

**Figure 3.**
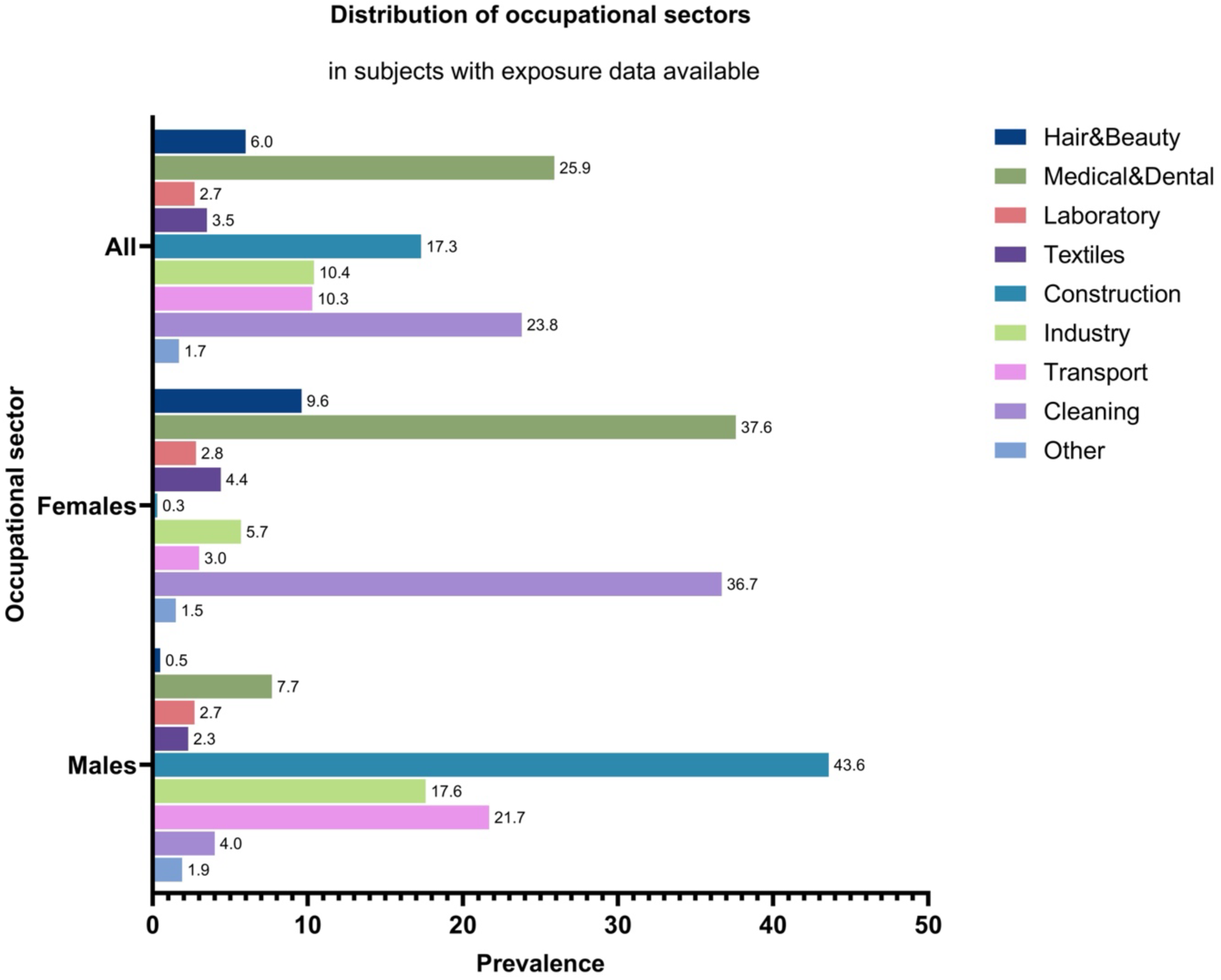
Distribution of occupational sectors among participants with occupational exposure to airborne MNP. Shown are the proportions of participants within a particular occupational sector of all subjects exposed to MNP: e.g. of all subjects exposed to MNP, 6.0% works in the Hair&Beauty sector.

### Correlations between occupational MNP exposure and other occupational exposures

As occupational exposures can co-occur, we also investigated correlations between MNP and other types of occupational exposures (Spearman rank correlations between MNP and other types of occupational exposures are presented in supplemental figure S3 to S5). Occupational MNP exposure was moderately correlated with occupational exposure to gasses and fumes (Rho=0.42) and solvents (Rho=0.48), and weakly correlated to occupational exposure to organic dust, pesticides, metals and silica. The correlation between MNP and gasses and fumes was larger in females (Rho=0.53) than in males (Rho=0.22) and the correlation between MNP and occupational exposure to silica was moderate in males only (Rho in men=0.56).

### Occupational MNP exposure is associated with lower lung function

To determine whether occupational MNP exposure is associated with respiratory health effects, we first examined lung function. In the base model, occupational MNP exposure was significantly associated with lower lung function and a higher prevalence of airway obstruction (supplementary table S8). After adjustment for co-exposures, i.e. the main model, participants with high occupational MNP exposure showed significantly lower lung function compared to those with no occupational exposure, with an average reduction of 42.9 ml in FEV₁, 47.4 ml in FVC and 0.26% in FEV_1_/FVC (Figure 4, supplementary table S9). High occupational MNP exposure was also associated with higher odds for airway obstruction. In both females and males, effect sizes were larger at higher exposure levels, although in males only the high exposure category reached statistical significance.

**Figure 4:**
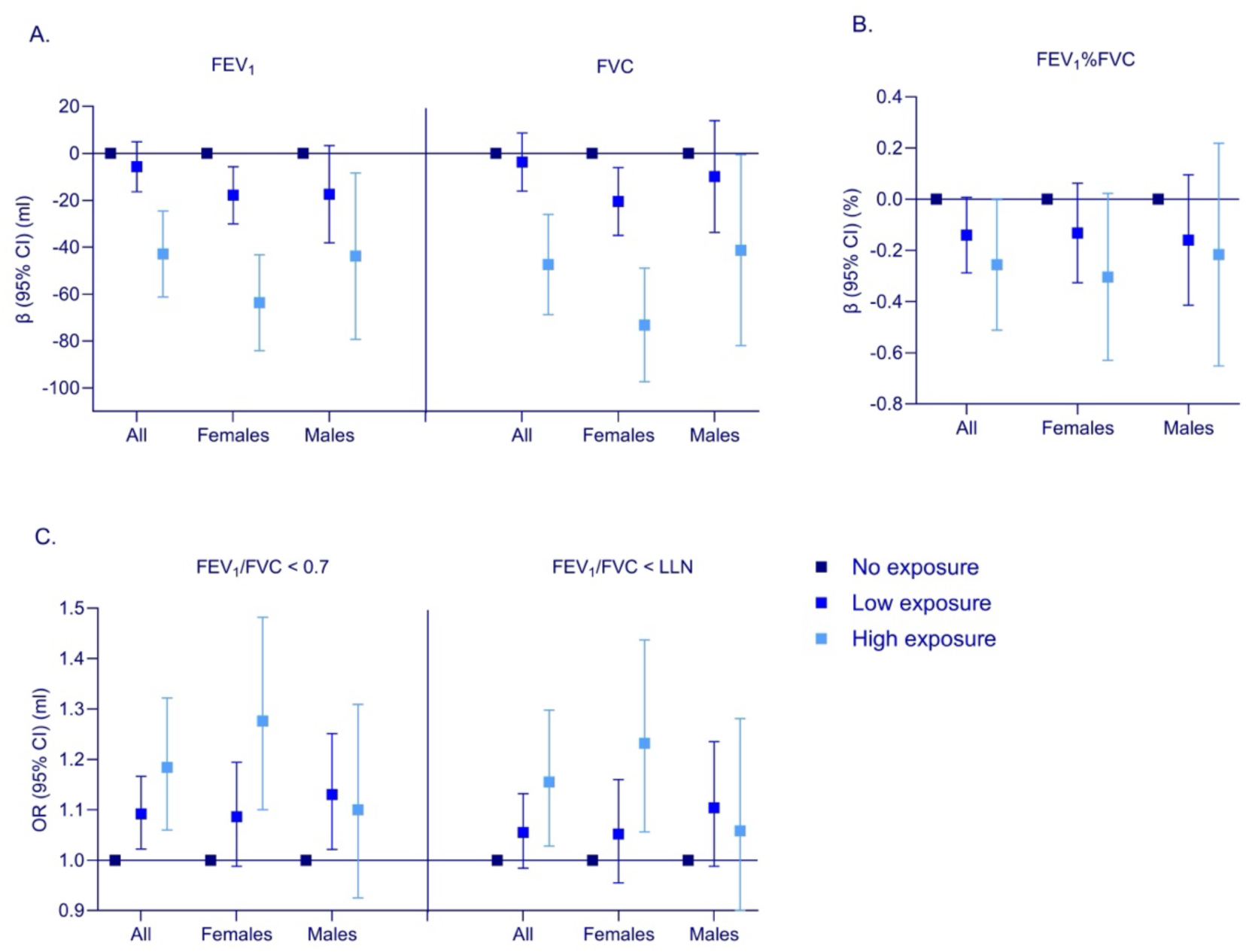
Associations between low and high occupational MNP exposure and lung function. Linear regression models with lung function as outcome (A & B), logistic regression models with airway obstruction as outcome (C). All models are adjusted for age, sex, height, smoking and occupational exposure to organic dust, gasses/fumes, pesticides, metals, solvents and silica (main model).

Sector-specific analyses showed that the overall associations were not driven by a single sector but were observed across multiple sectors with plausible MNP exposure, including Industry, Transport, Textiles and Cleaning (supplementary tables S10 and S11). These associations were similar in the sensitivity analysis with additional adjustment for education and income, although the effect estimates became smaller and were no longer significant in males (supplementary table S12).

### Occupational MNP exposure is associated with higher odds for respiratory symptoms and asthma

Figure 5 shows the results of the main model for associations between occupational exposure level and the presence of respiratory symptoms and asthma. Compared to those with no occupational exposure, participants with high exposure had a significantly higher odds for all outcomes except sputum. As with lung function, effect sizes were generally larger at higher exposure levels. In females, significant associations with occupational MNP exposure were found for cough, bronchitis and dyspnea, while in males wheeze and dyspnea were significantly associated (supplementary table S14). Slightly smaller effect estimates were observed in the sensitivity analyses with additional adjustment for education and income and only the association with dyspnea remained significant (supplementary table S15). In the base model (without adjustment for co-exposures and without adjustment for education and income), high occupational MNP exposure was significantly associated with a higher prevalence of all symptoms and asthma (supplementary table S13).

**Figure 5:**
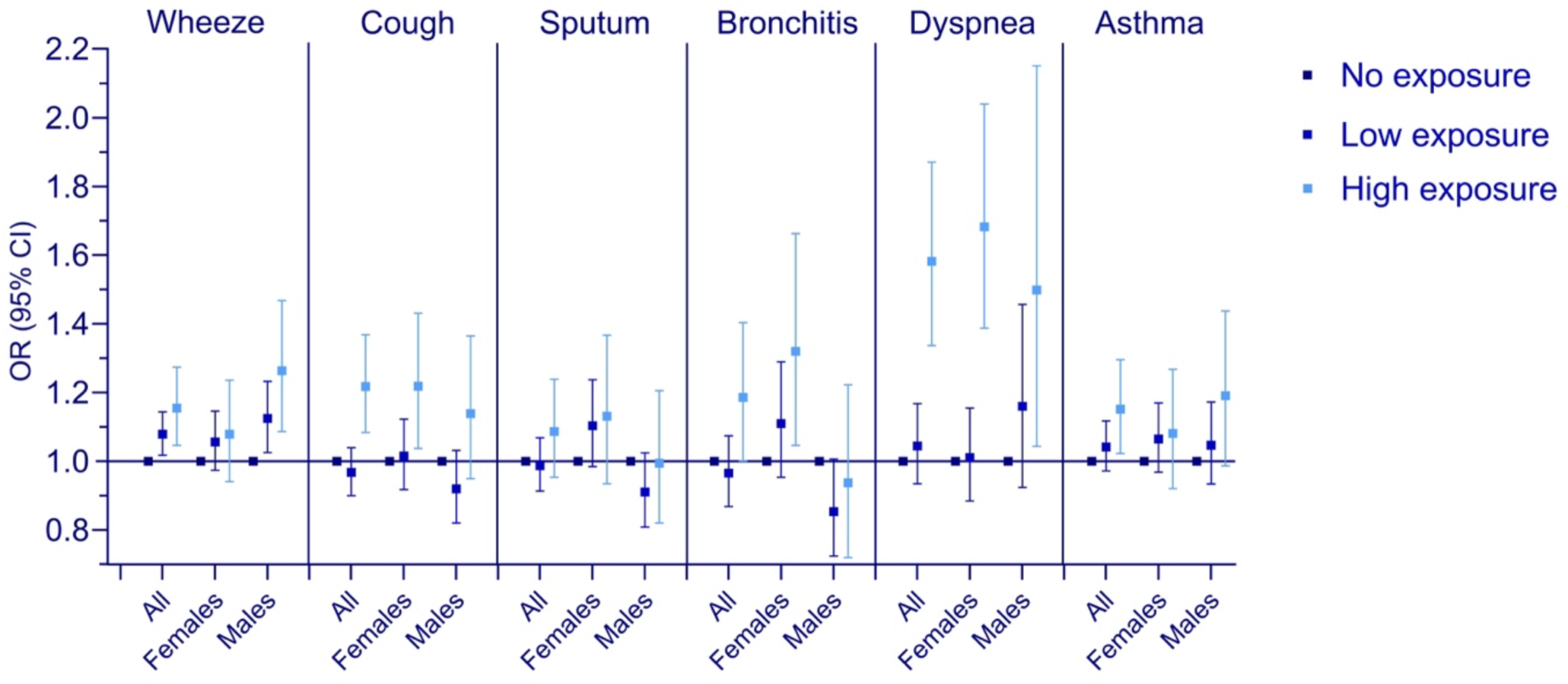
Associations between low and high occupational MNP exposure and respiratory symptoms and asthma. Logistic regression models adjusted for age, sex, BMI, smoking and occupational exposure to organic dust, gasses/fumes, pesticides, metals, solvents and silica (main model).

### No longitudinal association of occupational MNP exposure with respiratory outcomes

Beyond cross-sectional associations, we also investigated whether MNP exposure affects respiratory health over time. No significant associations were observed between occupational MNP exposure levels and annual decline in lung function (Table 2, supplementary table S16, S17), nor with the development of airway obstruction (Table 2, supplementary table S19, S20), respiratory symptoms or asthma (Table 2, supplementary table S22, S23). These findings were consistent in the sensitivity analyses (supplementary tables S18, S21 and S24).

**Table 2.** AssociaUons between occupaUonal MNP exposure and lung funcUon decline, development of airway obstrucUon and development of respiratory symptoms and asthma.

|  | n | estimate | 95% CI | P |
| --- | --- | --- | --- | --- |
| **Outcome: $\Delta$ FEV1 (ml/yr)** |  |  |  |  |
| No exposure | 34561 | reference |  |  |
| Low exposure | 9647 | 0.107 | -0.825;1.038 | 0.823 |
| High exposure | 1296 | -0.124 | -1.795;1.548 | 0.885 |
| **Outcome: $\Delta$ FVC (ml/yr)** |  |  |  |  |
| No exposure | 34561 | reference |  |  |
| Low exposure | 9647 | -0.331 | -1.539;0.876 | 0.591 |
| High exposure | 1296 | -0.626 | -2.793;1.541 | 0.571 |
| **Outcome: $\Delta$ FEV1%FVC (%/yr)** |  |  |  |  |
| No exposure | 34561 | reference |  |  |
| Low exposure | 9647 | 0.008 | -0.006;0.022 | 0.277 |
| High exposure | 1296 | 0.006 | -0.020;0.031 | 0.672 |

|  | n | OR | 95% CI | P |
| --- | --- | --- | --- | --- |
| **Outcome: development of FEV1/FVC < 0.7** |  |  |  |  |
| No exposure | 31268 | reference |  |  |
| Low exposure | 8514 | 1.026 | 0.905;1.163 | 0.686 |
| High exposure | 1106 | 1.032 | 0.824;1.294 | 0.782 |
| **Outcome: development of FEV1/FVC < LLN** |  |  |  |  |
| No exposure | 32184 | reference |  |  |
| Low exposure | 8832 | 0.979 | 0.823;1.164 | 0.808 |
| High exposure | 1152 | 0.842 | 0.608;1.166 | 0.842 |
| **Outcome: development of Wheeze** |  |  |  |  |
| No exposure | 47299 | reference |  |  |
| Low exposure | 13227 | 0.953 | 0.812;1.118 | 0.556 |
| High exposure | 1786 | 1.132 | 0.866;1.479 | 0.365 |
| **Outcome: development of Cough** |  |  |  |  |
| No exposure | 49716 | reference |  |  |
| Low exposure | 14031 | 0.965 | 0.863;1.079 | 0.537 |
| High exposure | 1883 | 1.132 | 0.941;1.362 | 0.188 |
| **Outcome: development of Sputum** |  |  |  |  |
| No exposure | 50337 | reference |  |  |
| Low exposure | 14165 | 0.970 | 0.858;1.096 | 0.622 |
| High exposure | 1922 | 0.976 | 0.786;1.211 | 0.824 |
| **Outcome: development of Bronchitis** |  |  |  |  |
| No exposure | 52008 | reference |  |  |
| Low exposure | 14673 | 0.976 | 0.846;1.126 | 0.736 |
| High exposure | 2007 | 0.948 | 0.736;1.221 | 0.679 |
| **Outcome: development of Asthma** |  |  |  |  |
| No exposure | 49343 | reference |  |  |
| Low exposure | 13870 | 1.020 | 0.886;1.174 | 0.783 |
| High exposure | 1905 | 1.142 | 0.901;1.448 | 0.273 |

Lung function decline: Linear mixed regression models with subject specific random intercept with lung function as outcome. The presented estimates are the coefficients from the interaction between time and MNP exposure at baseline. Restricted to subjects with at least two lung function measurements and $\geq 25$ years at baseline. Development of airway obstruction, respiratory symptoms and asthma: Logistic regression models. All models are adjusted for age, sex, height (outcome: lung function and airway obstruction) or BMI (outcome: symptoms/asthma), smoking, time since baseline (outcome: airway obstruction) and occupational exposure to organic dust, gasses/fumes, pesticides, metals, solvents and silica at baseline.

## Discussion

In this study, we developed a JEM for occupational exposure to airborne MNP (PlastiXJEM®) and applied it to the Lifelines cohort. Our findings suggest a dose-dependent association between occupational MNP exposure and lower lung function and higher prevalence of airway obstruction, respiratory symptoms and asthma, with larger effects at higher exposure levels. Importantly, the associations remained mostly significant after adjusting for education and income, which is consistent with an effect that is not fully explained by socio-economic status. However, no significant associations were found between MNP exposure and lung function decline over time or the development of airway obstruction, respiratory symptoms or asthma.

Our findings build upon emerging evidence that MNP exposure may negatively impact respiratory health. Previous epidemiological studies, primarily focusing on plastic manufacturing and processing workers, have consistently reported more respiratory morbidity, including higher risks for bronchial inflammation, lung cancer and interstitial lung diseases [13, 14, 17–24, 36–45]. Similarly, inhalation exposure to synthetic polymer dust among workers in plastic textile manufacturing settings has been linked to chronic respiratory symptoms and a lower lung function [20, 24, 46]. Our results align with and extend these findings by demonstrating similar associations in a general occupational context.

Although the observed effect sizes may appear modest at an individual level (e.g.,-43 ml for FEV₁), they are comparable to those reported for other established occupational exposures in population-based studies [47] and correspond to roughly 1-2 years of additional age-related lung function decline [48]. Moreover, these are population-level averages from a JEM-based study, where exposure misclassification is expected to attenuate effect estimates. The true effects in genuinely exposed individuals are therefore likely to be larger. The clinical relevance is further underscored by the fact that MNP exposure was associated not only with lower lung function but also with higher odds of airway obstruction, respiratory symptoms and asthma, indicating a consistent pattern across multiple respiratory outcomes.

In sex-stratified analyses, spirometry-defined airway obstruction was associated with MNP exposure in females only. For self-reported symptoms, associations differed by sex: in females, MNP exposure was associated with cough, bronchitis and dyspnea, whereas in males it was associated with wheeze and dyspnea. These sex-specific patterns can have several, not mutually exclusive, explanations.

First, males and females can receive a different effective dose of inhaled particles even at the same exposure level [49]. Differences in airway size and geometry, breathing patterns and lung volumes can affect where and how efficiently particles deposit along the respiratory tract, which may lead to higher airway deposition in females for some particle size fractions and therefore different symptoms. Second, respiratory symptoms are not interchangeable: they may point to different underlying processes. Cough and bronchitis-type symptoms are commonly linked to airway irritation and increased mucus [50], while wheeze is a hallmark of airway narrowing and variable airflow limitation [51]. Because airways of males and females can differ in size, inflammatory responses and symptom perception, an exposure that affects the airways may lead to a cough-predominant pattern in one sex and a wheeze-predominant pattern in the other [30, 31]. Third, even within the same occupational code, males and females may perform different tasks, work in different settings, or use different materials, leading to different co-exposure profiles and exposure intensity. In a JEM-based analysis, such within-code task differences can appear as sex-specific associations [52]. The observed differences between males and females should therefore be interpreted as potentially reflecting both biological susceptibility and differences in the exposure mixture and intensity across sectors and tasks.

Although we observed cross-sectional associations between occupational MNP exposure and lower lung function levels, no significant associations were found with the rate of lung function decline over time. This finding could have several different explanations. A lower lung function level does not necessarily imply an accelerated decline, as individuals may have reached a lower plateau earlier in life due to previous exposures or developmental factors affecting maximal lung growth [53]. Thus, exposed individuals could already have a lower baseline lung function without showing a faster decline during follow-up. This distinction between *lung function level* and *lung function decline* is well recognized and reflects two related but biologically distinct phenotypes of respiratory aging [47]. Furthermore, the follow-up period in Lifelines was relatively short and based on a limited number of spirometric measurements, which reduces statistical power to detect small differences in decline rates.

The estimated occupational MNP exposure in our newly developed JEM was moderately correlated with other occupational exposures as estimated with the OAsJEM and the EUROJEM. These correlations indicate that a substantial proportion of workers with higher MNP exposure are also exposed to other occupational agents, which complicates disentangling specific effects. Our analyses were adjusted for these co-exposures (i.e. the main model) which may have resulted in some overadjustment. In the base model, without adjustment for co-exposures, we observed consistent and statistically significant associations between MNP exposure and all cross-sectional lung function and symptom outcomes. Importantly, after additional adjustment for co-exposures, the majority of the associations remained significant. This pattern is compatible with an independent effect of MNP exposure on these outcomes that is not fully explained by other occupational exposures, although residual confounding and overadjustment cannot be excluded.

Several mechanisms could explain how microplastic exposure adversely impacts respiratory health. Experimental studies suggest that airborne MNP inhalation may cause direct mechanical irritation and damage to respiratory tissues, initiating inflammation and oxidative stress responses [10].

Chronic inflammation and sustained oxidative stress are known to impair lung function and contribute to symptoms such as cough and dyspnea [54]. Additionally, MNP contain plastic-associated additives and non-intentionally added substances [55], potentially exacerbating inflammation and tissue injury through chemical toxicity [55, 56]. Long-term exposure to these particles and associated chemicals may lead to chronic pulmonary inflammation, epithelial cell damage, and genomic instability, which are biological processes that could possibly contribute to an increased risk of respiratory diseases such as chronic obstructive pulmonary disease and lung cancer [57–59].

Despite these findings, several limitations of our study need consideration. First and foremost, the lack of direct measurement of airborne MNP exposure represents a significant limitation. Our JEM relied solely on expert judgment and literature-based inference regarding occupational exposure sources, such as synthetic textile fibers, cleaning agents, tire wear particles, and 3D printing materials. While such expert-based approaches are common for newly recognized occupational hazards lacking direct measurements, it introduces uncertainty and potential misclassification of exposure. In this context, it should be noted that concentrations of certain MNP sources, such as tire wear particles, are generally very low outdoors [60–62]. However, the PlastiXJEM®considered multiple potential sources beyond tire wear, including indoor occupational environments where synthetic face masks are routinely used [63–66], or where textile and synthetic materials are handled [8, 67]. Importantly, misclassification of exposure is likely to be largely nondifferential with respect to the outcomes and may bias associations towards the null, potentially underestimating true effects. In addition, because all subjects within the same ISCO code were assigned the same exposure category, the JEM is also expected to introduce a Berkson-type error [68], in which the assigned group-level exposure does not capture within-job variability. Such error generally reduces precision rather than causing systematic bias in the effect estimate. An additional source of potential underestimation is the healthy worker effect. Workers who develop respiratory problems may be more likely to leave jobs with high exposure or shift to less exposed occupations, meaning that the remaining workforce in high-exposure jobs may be healthier on average. This is particularly relevant for our cross-sectional analyses, where such selective job changes would weaken the observed associations.

Another limitation relates to the lack of complete occupational histories. In our study, exposure classification relied on the current or most recent occupation at baseline only. This means that cumulative lifetime exposures prior to baseline may be substantially underestimated, and that changes in occupation or exposure during the follow-up period are not captured. Consequently, cross-sectional associations should be interpreted as reflecting relatively recent exposure rather than cumulative lifetime exposure, and the ability to detect associations with lung function decline may therefore be reduced.

Despite these limitations, our study has notable strengths. It represents the first large-scale effort to systematically estimate occupational exposure to airborne MNP and investigate possible health impacts in a population-based cohort. The large sample size and standardised spirometry within the Lifelines cohort allowed us to examine a wide range of respiratory outcomes over time and explore potential sex-specific differences. Importantly, our analyses were adjusted not only for common confounders such as age, sex, smoking, height, and BMI, but also for occupational co-exposures to organic dust, gasses and fumes, pesticides, metals, solvents and silica using the OAsJEM and the EUROJEM. In addition, sensitivity analyses with adjustment for educational level and income showed that especially the observed associations with lung function largely persisted, suggesting that our findings are robust and not simply explained by socio-economic status or occupational co-exposures.

In conclusion, our findings provide evidence of an association between occupational exposure to airborne MNP and adverse respiratory health outcomes. Although some limitations regarding exposure estimation and the absence of detailed occupational histories highlight the preliminary nature of our results, this study underscores the clear need for future research using validated exposure measurement techniques and complete occupational data. Ultimately, these findings may provide a useful starting point for regulatory bodies to consider when developing occupational exposure guidelines. Moreover, because occupationally exposed workers represent the high end of the exposure spectrum, these results may also help inform the assessment of potential health risks from lower-level MNP exposure in everyday environments.

## Data Availability

The PlastiXJEM® (v1.0) is available to academic researchers for non-commercial use under specific access and use conditions. Requests should be directed to the corresponding author (Prof. Dr. Barbro N. Melgert). Lifelines individual-level data are not publicly available. Researchers can apply to access the Lifelines data used in this study via the Lifelines data access procedure; details and conditions are available on the Lifelines website.

## Acknowledgments

We thank Dr Pernilla Wiebert and Dr Håkan Tinnerberg (Karolinska Institutet and the EuroJEM chemical panel) for providing access to EuroJEM crystalline silica exposure.

## Declaration of interests

Barbro Melgert, Gwenda Vasse, Nienke Vrisekoop, and Joëlle Klazen report financial support was provided by Netherlands Organisation for Health Research and Development. Barbro Melgert has intellectual property #PlastiXJEM (job-exposure matrix for occupational airborne micro-and nanoplastics) deposited in the name of Barbro Melgert (BOIP i-DEPOT No. 157917). All authors declare that they have no known competing financial interests or personal relationships that could have appeared to influence the work reported in this manuscript.

## Author contributions

**Gwenda Vasse:** Conceptualization; Data curation; Investigation; Methodology; Project administration; Validation; Visualization; Writing – original draft; Writing – review & editing. **Nienke Vriesekoop:** Data curation; Methodology; Writing – review & editing.

**Joëlle Klazen:** Data curation; Methodology; Writing – review & editing.

**Judith Vonk:** Conceptualization; Data curation; Formal analysis; Investigation; Methodology; Project administration; Resources; Software; Supervision; Validation; Visualization; Writing – original draft; Writing – review & editing.

**Barbro Melgert:** Conceptualization; Data curation; Formal analysis; Funding acquisition; Investigation; Methodology; Project administration; Resources; Software; Supervision; Validation; Visualization; Writing – original draft; Writing – review & editing.

## Declaration of generative AI and AI-assisted technologies in the manuscript preparation process

During the preparation of this work the authors used Claude (Anthropic) for editing and grammar checking the text. After using this tool, the authors reviewed and edited the content as needed and take full responsibility for the content of the published article.

## Funding

This work is part of the MOMENTUM and MOMENTUM2.0 projects. The MOMENTUM project was made possible by ZonMw Programme Microplastics and Health, and Health-Holland, Top Sector Life Sciences & Health (project number 4580011101). The MOMENTUM2.0 project is funded by ZonMw Programme Microplastics and Health (project number 4580012310002). The Lifelines initiative has been made possible by subsidy from the Dutch Ministry of Health, Welfare and Sport, the Dutch Ministry of Economic Affairs, the University Medical Center Groningen (UMCG), University of Groningen and the Provinces in the North of the Netherlands (Drenthe, Friesland, Groningen)

## Supplemental tables

**Supplementary table S1.** Classification of the occupations with low or high MNP exposure into occupational sectors.

| Sector | ISCO-08 codes |
| --- | --- |
| Hair & Beauty | 5141, 5142 [1-5] |
| Medical & Dental | 2212, 2221, 2240, 2250, 2261, 3211, 3214, 3221, 3240, 3251, 3256, 3258 [6-17] |
| Laboratory | 2131, 2145, 2149, 3111, 3141, 3212 [6, 18-21] |
| Textiles | 2163, 7318, 7531, 7532, 7533, 7534, 7536, 8151, 8152, 8153, 8154, 8157, 8159, 9121 [22-30] |
| Construction | 3123, 7111, 7112, 7115, 7119, 7121, 7122, 7123, 7124, 7126, 7127, 7131, 7132, 7411, 7413, 7522, 7542, 9312, 9313, 9622 [31-36] |
| Industry | 3151, 7211, 7215, 7223, 7231, 7232, 7412, 7421, 7521, 8131, 8141, 8142, 8156, 8183, 8211, 8212, 8219, 9321, 9329, 9612 [37-43] |
| Transport | 3153, 5111, 8321, 8322, 8331, 8332, 8344, 9331 [12, 44-50] |
| Cleaning | 5151, 7133, 9111, 9112, 9129, 9611, 9613 [43, 51-56] |
| Other | 2651, 5245, 5411, 7316, 7541 [34, 57-61] |

**Supplementary table S2.** Definition of symptoms at baseline and at the first follow-up health assessment.

|  | Baseline | Follow-up |
| --- | --- | --- |
| **Wheeze** | Did you suffer from wheeze while you did not have a cold? | Have you suffered from wheeze in the past 12 months? |
| **Cough** | Do you usually cough in winter when getting up, during the daytime or at night on a daily basis for at least 3 months a year? | Do you usually cough on a daily basis for at least 3 months a year? |
| **Sputum** | Do you usually bring up sputum in winter when getting up, during the daytime or at night on a daily basis for at least 3 months a year? | Do you usually bring up sputum on a daily basis for at least 3 months a year? |
| **Bronchitis** | Do you usually cough and bring up sputum in winter when getting up, during the daytime or at night on a daily basis for at least 3 months a year? | Do you usually cough and bring up sputum on a daily basis for at least 3 months a year? |
| **Dyspnea** | Do you experience dyspnea when walking at normal pace with people of the same age on level ground? | NA |
| **Asthma** | Have you ever had a doctor's diagnosis of asthma? | Have you had an asthma attack in the past 12 months or do you currently use asthma medication? |

**Supplementary table S3.** Definition of covariates.

| **Covariate** | **Category** | **Definition** |
| --- | --- | --- |
| **Smoking** | Never | never smoked or smoked for <1 year |
|  | Ex | smoked for ≥1 year and stopped smoking for ≥1 month |
|  | Current | current smoker or stopped smoking <1 month |
|  | Unknown | inconsistent reporting or missing |
| **Education** | Low | no training, primary education, lower or prevocational education |
|  | Medium | general secondary education, secondary vocational or professional guiding, preuniversity education |
|  | High | higher professional or university degree |
|  | Unclassifiable | other than above-mentioned education or missing data |
| **Income** | Low | monthly net income ≤ €1500 |
|  | Medium | monthly net income between €1500 up and €2500 |
|  | High | monthly net income ≥ €2500 |
|  | Unknown | I do not know/I do not want to say/missing |

**Supplementary table S4.** Descriptives statistics stratified by MNP exposure for subjects included in the analysis on lung function level – Total group and stratified by sex.

|  | Total group |  |  |  | Females |  |  |  | Males |  |  |  |
| --- | --- | --- | --- | --- | --- | --- | --- | --- | --- | --- | --- | --- |
|  | All | No exposure | Low exposure | High exposure | All Females | No exposure | Low exposure | High exposure | All Males | No exposure | Low exposure | High exposure |
| **N** | 94844 | 71358 (75.2) | 20453 (21.6) | 3033 (3.2) | 58275 | 43951 (75.4) | 12748 (21.9) | 1576 (2.7) | 36569 | 27407 (74.9) | 7705 (21.1) | 1457 (4.0) |
| **Age, year** | 44.2 (12.7) | 44.0 (12.7) | 45.0 (12.6) | 45.3 (12.7) | 43.9 (12.5) | 43.5 (12.5) | 45.2 (12.6) | 45.7 (13.0) | 44.8 (12.9) | 44.9 (13.0) | 44.6 (12.5) | 44.8 (12.4) |
| **Male sex** | 36569 (38.6) | 27407 (38.4) | 7705 (37.7) | 1457 (48.0) |  |  |  |  |  |  |  |  |
| **Height, cm** | 174.4 (9.3) | 174.6 (9.3) | 173.7 (9.3) | 174.4 (9.6) | 169.3 (6.5) | 169.5 (6.5) | 168.8 (6.5) | 168.2 (7.1) | 182.5 (7.0) | 182.8 (7.0) | 181.9 (7.1) | 181.2 (7.1) |
| **Weight, kg** | 79.3 (15.1) | 79.1 (15.0) | 79.7 (15.5) | 81.2 (15.9) | 74.0 (13.8) | 73.9 (13.7) | 74.4 (13.9) | 75.3 (15.7) | 87.7 (13.2) | 87.5 (13.0) | 88.6 (13.9) | 87.6 (13.5) |
| **BMI, kg/m<sup>2</sup>** | 26.0 (4.3) | 25.9 (4.2) | 26.4 (4.5) | 26.7 (4.7) | 25.8 (4.7) | 25.7 (4.6) | 26.1 (4.8) | 26.7 (5.4) | 26.3 (3.6) | 26.2 (3.6) | 26.8 (3.9) | 26.7 (3.7) |
| **Smoking** |  |  |  |  |  |  |  |  |  |  |  |  |
| **Never** | 42051 (44.3) | 32355 (45.3) | 8520 (41.7) | 1176 (38.8) | 26824 (46.0) | 20424 (46.5) | 5696 (44.7) | 704 (44.7) | 15227 (41.6) | 11931 (43.5) | 2824 (36.7) | 472 (32.4) |
| **Ex** | 30043 (31.7) | 22384 (31.4) | 6737 (32.9) | 922 (30.4) | 18196 (31.2) | 13550 (30.8) | 4202 (33.0) | 444 (28.2) | 11847 (32.4) | 8834 (32.2) | 2535 (32.9) | 478 (32.8) |
| **Current** | 18949 (20.0) | 13717 (19.2) | 4437 (21.7) | 795 (26.2) | 10911 (18.7) | 8146 (18.5) | 2398 (18.8) | 367 (23.3) | 8038 (22.0) | 5571 (20.3) | 2039 (26.5) | 428 (29.4) |
| **Missing** | 3801 (4.0) | 2902 (4.1) | 759 (3.7) | 140 (4.6) | 2344 (4.0) | 1831 (4.2) | 452 (3.5) | 61 (3.9) | 1457 (4.0) | 1071 (3.9) | 307 (4.0) | 79 (5.4) |
| **Education** |  |  |  |  |  |  |  |  |  |  |  |  |
| **Low** | 28384 (29.9) | 17726 (24.8) | 8851 (43.3) | 1807 (59.6) | 17207 (29.5) | 11357 (25.8) | 4973 (39.0) | 877 (55.6) | 11177 (30.6) | 6369 (23.2) | 3878 (50.3) | 930 (63.8) |
| **Medium** | 37395 (39.4) | 28925 (40.5) | 7556 (36.9) | 914 (30.1) | 23749 (40.8) | 18444 (42.0) | 4782 (37.5) | 523 (33.2) | 13646 (37.3) | 10481 (38.2) | 2774 (36.0) | 391 (26.8) |
| **High** | 27406 (28.9) | 23519 (33.0) | 3643 (17.8) | 244 (8.0) | 16293 (28.0) | 13420 (30.5) | 2729 (21.4) | 144 (9.1) | 11113 (30.4) | 10099 (36.8) | 914 (11.9) | 100 (6.9) |
| **Missing** | 1659 (1.7) | 1188 (1.7) | 403 (2.0) | 68 (2.2) | 1026 (1.8) | 730 (1.7) | 264 (2.1) | 32 (2.0) | 633 (1.7) | 458 (1.7) | 139 (1.8) | 36 (2.5) |
| **Income** |  |  |  |  |  |  |  |  |  |  |  |  |
| **Low** | 14260 (15.0) | 10274 (14.4) | 3370 (16.5) | 616 (20.3) | 10009 (17.2) | 7401 (16.8) | 2245 (17.6) | 363 (23.0) | 4251 (11.6) | 2873 (10.5) | 1125 (14.6) | 253 (17.4) |
| **Medium** | 24587 (25.9) | 17347 (24.3) | 6361 (31.1) | 879 (29.0) | 14282 (24.5) | 10333 (23.5) | 3555 (27.9) | 394 (25.0) | 10305 (28.2) | 7014 (25.6) | 2806 (36.4) | 485 (33.3) |
| **High** | 39642 (41.8) | 32067 (44.9) | 6774 (33.1) | 801 (26.4) | 23078 (39.6) | 18432 (41.9) | 4247 (33.3) | 399 (25.3) | 16564 (45.3) | 13635 (49.8) | 2527 (32.8) | 402 (27.6) |
| **Unknown** | 16355 (17.2) | 11670 (16.4) | 3948 (19.3) | 737 (24.3) | 10906 (18.7) | 7785 (17.7) | 2701 (21.2) | 420 (26.6) | 5449 (14.9) | 3885 (14.2) | 1247 (16.2) | 317 (21.8) |
| **Organic dust exposure<sup>a</sup>** |  |  |  |  |  |  |  |  |  |  |  |  |
| **No** | 87646 (92.4) | 61744 (94.1) | 17848 (87.3) | 2654 (87.5) | 56670 (97.2) | 42660 (97.1) | 12468 (97.8) | 1542 (97.8) | 30976 (84.7) | 24484 (89.3) | 5380 (69.8) | 1112 (76.3) |
| **Low** | 3592 (3.8) | 1214 (1.7) | 2079 (10.2) | 299 (9.9) | 503 (0.9) | 414 (0.9) | 66 (0.5) | 23 (1.5) | 3089 (8.4) | 800 (2.9) | 2013 (26.1) | 276 (18.9) |
| **High** | 3606 (3.8) | 3000 (4.2) | 526 (2.6) | 80 (2.6) | 1102 (1.9) | 877 (2.0) | 214 (1.7) | 11 (0.7) | 2504 (6.8) | 2123 (7.7) | 312 (4.0) | 69 (4.7) |
| **Gasses/Fumes exposure<sup>a</sup>** |  |  |  |  |  |  |  |  |  |  |  |  |
| **No** | 68317 (72.0) | 59880 (83.9) | 6290 (30.8) | 2147 (70.8) | 40867 (70.1) | 37715 (85.8) | 2251 (17.7) | 901 (57.2) | 27450 (75.1) | 22165 (80.9) | 4039 (52.4) | 1246 (85.5) |
| **Low** | 14740 (15.5) | 5066 (7.1) | 8902 (43.5) | 772 (25.5) | 9790 (16.8) | 1884 (4.3) | 7277 (57.1) | 629 (39.9) | 4950 (13.5) | 3182 (11.6) | 1625 (21.1) | 143 (9.8) |
| **High** | 11787 (12.4) | 6412 (9.0) | 5261 (25.7) | 114 (3.8) | 7618 (13.1) | 4352 (9.9) | 3220 (25.3) | 46 (2.9) | 4169 (11.4) | 2060 (7.5) | 2041 (26.5) | 68 (4.7) |
| **Pesticide exposure<sup>a</sup>** |  |  |  |  |  |  |  |  |  |  |  |  |
| **No** | 90409 (95.3) | 67300 (94.3) | 20128 (98.4) | 2981 (98.3) | 56918 (97.7) | 42705 (97.2) | 12654 (99.3) | 1559 (98.9) | 33491 (91.6) | 24595 (89.7) | 7474 (97.0) | 1422 (97.6) |
| **Low** | 2771 (2.9) | 2536 (3.6) | 215 (1.1) | 20 (0.7) | 846 (1.5) | 786 (1.8) | 54 (0.4) | 6 (0.4) | 1925 (5.3) | 1750 (6.4) | 161 (2.1) | 14 (1.0) |
| **High** | 1664 (1.8) | 1522 (2.1) | 110 (0.5) | 32 (1.1) | 511 (0.9) | 460 (1.0) | 40 (0.3) | 11 (0.7) | 1153 (3.2) | 1062 (3.9) | 70 (0.9) | 21 (1.4) |
| **Metals exposure<sup>a</sup>** |  |  |  |  |  |  |  |  |  |  |  |  |
| **No** | 90938 (95.9) | 69582 (97.5) | 18517 (90.5) | 2839 (93.6) | 57986 (99.5) | 43912 (99.9) | 12539 (98.4) | 1535 (97.4) | 32952 (90.1) | 25670 (93.7) | 5978 (77.6) | 1304 (89.5) |
| **Low** | 3092 (3.3) | 1265 (1.8) | 1647 (8.1) | 180 (5.9) | 278 (0.5) | 32 (0.1) | 205 (1.6) | 41 (2.6) | 2814 (7.7) | 1233 (4.5) | 1442 (18.7) | 139 (9.5) |
| **High** | 814 (0.9) | 511 (0.7) | 289 (1.4) | 14 (0.5) | 11 (0.0) | 7 (0.0) | 4 (0.0) | 0 (0.0) | 803 (2.2) | 504 (1.8) | 285 (3.7) | 14 (1.0) |
| **Solvents exposure<sup>a</sup>** |  |  |  |  |  |  |  |  |  |  |  |  |
| **No** | 81711 (86.2) | 68310 (95.7) | 11530 (56.4) | 1871 (61.7) | 51424 (88.2) | 42643 (97.0) | 7489 (58.7) | 1292 (82.0) | 30287 (82.8) | 25667 (93.7) | 4041 (52.4) | 579 (39.7) |
| **Low** | 10788 (11.4) | 2780 (3.9) | 7559 (37.0) | 449 (14.8) | 5313 (9.1) | 1234 (2.8) | 3944 (30.9) | 135 (8.6) | 5475 (15.0) | 1546 (5.6) | 3615 (46.9) | 314 (21.6) |
| **High** | 2345 (2.5) | 268 (0.4) | 1364 (6.7) | 713 (23.5) | 1538 (2.6) | 74 (0.2) | 1315 (10.3) | 149 (9.5) | 807 (2.2) | 194 (0.7) | 49 (0.6) | 564 (38.7) |
| **Silica exposure<sup>b</sup>** |  |  |  |  |  |  |  |  |  |  |  |  |
| **No** | 88613 (93.4) | 70026 (98.1) | 16325 (79.8) | 2262 (74.6) | 57873 (99.3) | 43670 (99.4) | 12656 (99.3) | 1547 (98.2) | 30740 (84.1) | 26356 (96.2) | 3669 (47.6) | 715 (49.1) |
| **Low** | 2490 (2.6) | 325 (0.5) | 1797 (8.8) | 368 (12.1) | 131 (0.2) | 66 (0.2) | 53 (0.4) | 12 (0.8) | 2359 (6.5) | 259 (0.9) | 1744 (22.6) | 356 (24.4) |
| **High** | 3741 (3.9) | 1007 (1.4) | 2331 (11.4) | 403 (13.3) | 271 (0.5) | 215 (0.5) | 39 (0.3) | 17 (1.1) | 3470 (9.5) | 792 (2.9) | 2292 (29.7) | 386 (26.5) |
| **Lung function** |  |  |  |  |  |  |  |  |  |  |  |  |
| **FEV1, L** | 3.45 (0.8) | 3.47 (0.8) | 3.39 (0.8) | 3.42 (0.9) | 3.03 (0.6) | 3.05 (0.6) | 2.97 (0.6) | 2.89 (0.6) | 4.12 (0.8) | 4.14 (0.8) | 4.07 (0.8) | 4.00 (0.8) |
| **FVC, L** | 4.49 (1.0) | 4.51 (1.0) | 4.42 (1.0) | 4.49 (1.1) | 3.91 (0.6) | 3.93 (0.6) | 3.86 (0.6) | 3.76 (0.7) | 5.41 (0.9) | 5.43 (0.9) | 5.36 (0.9) | 5.28 (0.9) |
| **FEV1%FVC, %** | 76.9 (7.2) | 77.1 (7.2) | 76.6 (7.3) | 76.3 (7.7) | 77.5 (7.0) | 77.6 (7.0) | 77.0 (7.1) | 76.8 (7.7) | 76.1 (7.5) | 76.2 (7.5) | 75.8 (7.6) | 75.7 (7.7) |
| **FEV1/FVC < 0.7** | 13847 (14.6) | 10117 (14.2) | 3201 (15.7) | 529 (17.4) | 7366 (12.6) | 5347 (12.2) | 1761 (13.8) | 258 (16.4) | 6481 (17.7) | 4770 (17.4) | 1440 (18.7) | 271 (18.6) |
| **FEV1/FVC < LLN** | 11243 (11.9) | 8257 (11.6) | 2564 (12.5) | 422 (13.9) | 6701 (11.5) | 4967 (11.3) | 1513 (11.9) | 221 (14.0) | 4542 (12.4) | 3290 (12.0) | 1051 (13.6) | 201 (13.8) |
| **Occupational Sectors\*** |  |  |  |  |  |  |  |  |  |  |  |  |
| **Hair&beauty** |  |  | 1356 (6.6) | 40 (1.3) |  |  | 1313 (10.3) | 40 (2.5) |  |  | 43 (0.6) | 0 (0.0) |
| **Medical/dental** |  |  | 5640 (27.6) | 611 (20.1) |  |  | 5037 (39.5) | 502 (31.9) |  |  | 603 (7.8) | 109 (7.5) |
| **Laboratory** |  |  | 682 (3.3) | 1 (0.0) |  |  | 415 (3.3) | 0 (0.0) |  |  | 267 (3.5) | 1 (0.1) |
| **Textiles** |  |  | 58 (0.3) | 766 (25.3) |  |  | 26 (0.2) | 601 (38.1) |  |  | 32 (0.4) | 165 (11.3) |
| **Construction** |  |  | 3170 (15.5) | 876 (28.9) |  |  | 22 (0.2) | 17 (1.1) |  |  | 3148 (40.9) | 859 (59.0) |
| **Industry** |  |  | 1673 (8.2) | 720 (23.7) |  |  | 361 (2.8) | 424 (26.9) |  |  | 1312 (17.0) | 296 (20.3) |
| **Transport** |  |  | 2376 (11.6) | 28 (0.9) |  |  | 422 (3.3) | 5 (0.3) |  |  | 1954 (25.4) | 23 (1.6) |
| **Cleaning** |  |  | 5340 (26.1) | 142 (4.7) |  |  | 5052 (39.6) | 80 (5.1) |  |  | 288 (3.7) | 62 (4.3) |
| **Other** |  |  | 405 (2.0) | 5 (0.2) |  |  | 229 (1.8) | 3 (0.2) |  |  | 176 (2.3) | 2 (0.1) |

Mean (SD) for continuous variables, N (%) for categorical variables. \* Occupational sectors: The numbers do not add up to the total for each exposure group since some subjects have more than 1 job with MNP exposure. <sup>a</sup> Exposures were based on the OAsJEM [62]: organic dust=highest level of animals, flour, foods, plants-related dusts, moulds, endotoxin; gasses/fumes=highest level of exhaust fumes, high-level chemical disinfectants, bleach; pesticides=highest level of herbicides, insecticides, fungicides; metals=highest level of metal, metal working fluids; solvents=organic solvents. <sup>b</sup> Exposure is based on the EUROJEM respirable crystalline silica [63].

**Supplementary table S5.** Descriptives statistics stratified by MNP exposure for subjects included in the analysis on respiratory symptoms and asthma – Total group and stratified by sex.

|  | Total group |  |  |  | Females |  |  |  | Males |  |  |  |
| --- | --- | --- | --- | --- | --- | --- | --- | --- | --- | --- | --- | --- |
|  | All | No exposure | Low exposure | High exposure | All Females | No exposure | Low exposure | High exposure | All Males | No exposure | Low exposure | High exposure |
| **N** | 133407 | 100662 (75.5) | 28645 (21.5) | 4100 (3.1) | 81641 | 61783 (75.7) | 17783 (21.8) | 2075 (2.5) | 51766 | 38879 (75.1) | 10862 (21.0) | 2025 (3.9) |
| **Age, yr** | 44.2 (12.9) | 44.0 (13.0) | 44.9 (12.7) | 44.9 (12.4) | 43.8 (12.8) | 43.4 (12.8) | 45.0 (12.7) | 45.2 (12.8) | 44.9 (13.1) | 45.0 (13.3) | 44.7 (12.7) | 44.6 (12.0) |
| **Male sex** | 51766 (38.8) | 38879 (38.6) | 10862 (37.9) | 2025 (49.4) |  |  |  |  |  |  |  |  |
| **Height, cm** | 174.5 (9.3) | 174.7 (9.3) | 173.9 (9.2) | 174.9 (9.6) | 169.4 (6.5) | 169.6 (6.5) | 168.9 (6.5) | 168.3 (6.8) | 182.6 (7.0) | 182.9 (7.0) | 182.0 (7.0) | 181.6 (7.0) |
| **Weight, kg** | 79.3 (15.3) | 79.1 (15.1) | 79.8 (15.7) | 81.5 (15.9) | 74.0 (13.9) | 73.8 (13.8) | 74.4 (14.1) | 75.1 (15.3) | 87.8 (13.4) | 87.5 (13.2) | 88.6 (14.0) | 88.0 (13.8) |
| **BMI, kg/m<sup>2</sup>** | 26.0 (4.4) | 25.9 (4.3) | 26.3 (4.5) | 26.6 (4.6) | 25.8 (4.7) | 25.7 (4.7) | 26.1 (4.9) | 26.5 (5.3) | 26.3 (3.7) | 26.2 (3.6) | 26.7 (3.9) | 26.7 (3.8) |
| **Smoking** |  |  |  |  |  |  |  |  |  |  |  |  |
| **Never** | 59168 (44.4) | 45688 (45.4) | 11887 (41.5) | 1593 (38.9) | 37652 (46.1) | 28820 (46.6) | 7912 (44.5) | 920 (44.3) | 21516 (41.6) | 16868 (43.4) | 3975 (36.6) | 673 (33.2) |
| **Ex** | 41818 (31.3) | 31311 (31.1) | 9296 (32.5) | 1211 (29.5) | 25157 (30.8) | 18830 (30.5) | 5753 (32.4) | 574 (27.7) | 16661 (32.2) | 12481 (32.1) | 3543 (32.6) | 637 (31.5) |
| **Current** | 26529 (19.9) | 19155 (19.0) | 6283 (21.9) | 1091 (26.6) | 15155 (18.6) | 11254 (18.2) | 3419 (19.2) | 482 (23.2) | 11374 (22.0) | 7901 (20.3) | 2864 (26.4) | 609 (30.1) |
| **Missing** | 5892 (4.4) | 4508 (4.5) | 1179 (4.1) | 205 (5.0) | 3677 (4.5) | 2879 (4.7) | 699 (3.9) | 99 (4.8) | 2215 (4.3) | 1629 (4.2) | 480 (4.4) | 106 (5.2) |
| **Education** |  |  |  |  |  |  |  |  |  |  |  |  |
| **Low** | 39497 (29.6) | 24767 (24.6) | 12343 (43.1) | 2387 (58.2) | 23908 (29.3) | 15922 (25.8) | 6863 (38.6) | 1123 (54.1) | 15589 (30.1) | 8845 (22.8) | 5480 (50.5) | 1264 (62.4) |
| **Medium** | 52789 (39.6) | 40878 (40.6) | 10624 (37.1) | 1287 (31.4) | 33388 (40.9) | 25939 (42.0) | 6736 (37.9) | 713 (34.4) | 19401 (37.5) | 14939 (38.4) | 3888 (35.8) | 574 (28.3) |
| **High** | 38774 (29.1) | 33324 (33.1) | 5112 (17.8) | 338 (8.2) | 22946 (28.1) | 18929 (30.6) | 3821 (21.5) | 196 (9.4) | 15828 (30.6) | 14395 (37.0) | 1291 (11.9) | 142 (7.0) |
| **Missing** | 2347 (1.8) | 1693 (1.7) | 566 (2.0) | 88 (2.1) | 1399 (1.7) | 993 (1.6) | 363 (2.0) | 43 (2.1) | 948 (1.8) | 700 (1.8) | 203 (1.9) | 45 (2.2) |
| **Income** |  |  |  |  |  |  |  |  |  |  |  |  |
| **Low** | 21224 (15.9) | 15335 (15.2) | 5009 (17.5) | 880 (21.5) | 14932 (18.3) | 11046 (17.9) | 3375 (19.0) | 511 (24.6) | 6292 (12.2) | 4289 (11.0) | 1634 (15.0) | 369 (18.2) |
| **Medium** | 34798 (26.1) | 24542 (24.4) | 9010 (31.5) | 1246 (30.4) | 19999 (24.5) | 14488 (23.4) | 4963 (27.9) | 548 (26.4) | 14799 (28.6) | 10054 (25.9) | 4047 (37.3) | 698 (34.5) |
| **High** | 56084 (42.0) | 45361 (45.1) | 9585 (33.5) | 1138 (27.8) | 32309 (39.6) | 25772 (41.7) | 5987 (33.7) | 550 (26.5) | 23775 (45.9) | 19589 (50.4) | 3598 (33.1) | 588 (29.0) |
| **Unknown** | 21301 (16.0) | 15424 (15.3) | 5041 (17.6) | 836 (20.4) | 14401 (17.6) | 10477 (17.0) | 3458 (19.4) | 466 (22.5) | 6900 (13.3) | 4947 (12.7) | 1583 (14.6) | 370 (18.3) |
| **Organic dust exposure<sup>a</sup>** |  |  |  |  |  |  |  |  |  |  |  |  |
| **No** | 123391 (92.5) | 94797 (94.2) | 25042 (87.4) | 3552 (86.6) | 79432 (97.3) | 59991 (97.1) | 17411 (97.9) | 2030 (97.8) | 43959 (84.9) | 34806 (89.5) | 7631 (70.3) | 1522 (75.2) |
| **Low** | 5019 (3.8) | 1704 (1.7) | 2887 (10.1) | 428 (10.4) | 666 (0.8) | 556 (0.9) | 83 (0.5) | 27 (1.3) | 4353 (8.4) | 1148 (3.0) | 2804 (25.8) | 401 (19.8) |
| **High** | 4997 (3.7) | 4161 (4.1) | 716 (2.5) | 120 (2.9) | 1543 (1.9) | 1236 (2.0) | 289 (1.6) | 18 (0.9) | 3454 (6.7) | 2925 (7.5) | 427 (3.9) | 102 (5.0) |
| **Gasses/Fumes exposure<sup>a</sup>** |  |  |  |  |  |  |  |  |  |  |  |  |
| **No** | 96156 (72.1) | 84418 (83.9) | 8852 (30.9) | 2886 (70.4) | 57262 (70.1) | 52933 (85.7) | 3163 (17.8) | 1166 (56.2) | 38894 (75.1) | 31485 (81.0) | 5689 (52.4) | 1720 (84.9) |
| **Low** | 20820 (15.6) | 7283 (7.2) | 12482 (43.6) | 1055 (25.7) | 13762 (16.9) | 2757 (4.5) | 10156 (57.1) | 849 (40.9) | 7058 (13.6) | 4526 (11.6) | 2326 (21.4) | 206 (10.2) |
| **High** | 16431 (12.3) | 8961 (8.9) | 7311 (25.5) | 159 (3.9) | 10617 (13.0) | 6093 (9.9) | 4464 (25.1) | 60 (2.9) | 5814 (11.2) | 2868 (7.4) | 2847 (26.2) | 99 (4.9) |
| **Pesticide exposure<sup>a</sup>** |  |  |  |  |  |  |  |  |  |  |  |  |
| **No** | 127037 (95.2) | 94820 (94.2) | 28194 (98.4) | 4023 (98.1) | 79651 (97.6) | 59940 (97.0) | 17660 (99.3) | 2051 (98.8) | 47386 (91.5) | 34880 (89.7) | 10534 (97.0) | 1972 (97.4) |
| **Low** | 4051 (3.0) | 3711 (3.7) | 309 (1.1) | 31 (0.8) | 1287 (1.6) | 1201 (1.9) | 75 (0.4) | 11 (0.5) | 2764 (5.3) | 2510 (6.5) | 234 (2.2) | 20 (1.0) |
| **High** | 2319 (1.7) | 2131 (2.1) | 142 (0.5) | 46 (1.1) | 703 (0.9) | 642 (1.0) | 48 (0.3) | 13 (0.6) | 1616 (3.1) | 1489 (3.8) | 94 (0.9) | 33 (1.6) |
| **Metals exposure<sup>a</sup>** |  |  |  |  |  |  |  |  |  |  |  |  |
| **No** | 127979 (95.9) | 98174 (97.5) | 25957 (90.6) | 3848 (93.9) | 81262 (99.5) | 61722 (99.9) | 17517 (98.5) | 2023 (97.5) | 46717 (90.2) | 36452 (93.8) | 8440 (77.7) | 1825 (90.1) |
| **Low** | 4318 (3.2) | 1798 (1.8) | 2284 (8.0) | 236 (5.8) | 359 (0.4) | 49 (0.1) | 258 (1.5) | 52 (2.5) | 3959 (7.6) | 1749 (4.5) | 2026 (18.7) | 184 (9.1) |
| **High** | 1110 (0.8) | 690 (0.7) | 404 (1.4) | 16 (0.4) | 20 (0.0) | 12 (0.0) | 8 (0.0) | 0 (0.0) | 1090 (2.1) | 678 (1.7) | 396 (3.6) | 16 (0.8) |
| **Solvents exposure<sup>a</sup>** |  |  |  |  |  |  |  |  |  |  |  |  |
| **No** | 115051 (86.2) | 96325 (95.7) | 16167 (56.4) | 2559 (62.4) | 72089 (88.3) | 59938 (97.0) | 10446 (58.7) | 1705 (82.2) | 42962 (83.0) | 36387 (93.6) | 5721 (52.7) | 854 (42.2) |
| **Low** | 15071 (11.3) | 3956 (3.9) | 10517 (36.7) | 598 (14.6) | 7373 (9.0) | 1752 (2.8) | 5446 (30.6) | 175 (8.4) | 7698 (14.9) | 2204 (5.7) | 5071 (46.7) | 423 (20.9) |
| **High** | 3285 (2.5) | 381 (0.4) | 1961 (6.8) | 943 (23.0) | 2179 (2.7) | 93 (0.2) | 1891 (10.6) | 195 (9.4) | 1106 (2.1) | 288 (0.7) | 70 (0.6) | 748 (36.9) |
| **Silica exposure<sup>b</sup>** |  |  |  |  |  |  |  |  |  |  |  |  |
| **No** | 124685 (93.5) | 98806 (98.2) | 22839 (79.7) | 3040 (74.1) | 81085 (99.3) | 61389 (99.4) | 17658 (99.3) | 2038 (98.2) | 43600 (84.2) | 37417 (96.2) | 5181 (47.7) | 1002 (49.5) |
| **Low** | 3503 (2.6) | 460 (0.5) | 2542 (8.9) | 501 (12.2) | 183 (0.2) | 95 (0.2) | 72 (0.4) | 16 (0.8) | 3320 (6.4) | 365 (0.9) | 2470 (22.7) | 485 (24.0) |
| **High** | 5219 (3.9) | 1396 (1.4) | 3264 (11.4) | 559 (13.6) | 373 (0.5) | 299 (0.5) | 53 (0.3) | 21 (1.0) | 4846 (9.4) | 1097 (2.8) | 3211 (29.6) | 538 (26.6) |
| **Respiratory symptoms** |  |  |  |  |  |  |  |  |  |  |  |  |
| **Wheeze** | 16441 (12.4) | 12077 (12.0) | 3767 (13.2) | 597 (14.7) | 9695 (11.9) | 7224 (11.7) | 2196 (12.4) | 275 (13.3) | 6746 (13.1) | 4853 (12.5) | 1571 (14.6) | 322 (16.1) |
| **Cough** | 10043 (7.6) | 7382 (7.4) | 2245 (7.9) | 416 (10.2) | 5871 (7.2) | 4387 (7.1) | 1291 (7.3) | 193 (9.4) | 4172 (8.1) | 2995 (7.8) | 954 (8.8) | 223 (11.1) |
| **Sputum** | 8255 (6.2) | 6023 (6.0) | 1912 (6.7) | 320 (7.9) | 4396 (5.4) | 3253 (5.3) | 1014 (5.7) | 129 (6.3) | 3859 (7.5) | 2770 (7.2) | 898 (8.3) | 191 (9.5) |
| **Bronchitis** | 4347 (3.3) | 3164 (3.1) | 997 (3.5) | 186 (4.5) | 2444 (3.0) | 1804 (2.9) | 554 (3.1) | 86 (4.2) | 1903 (3.7) | 1360 (3.5) | 443 (4.1) | 100 (4.9) |
| **Dyspnea** | 4041 (3.4) | 2883 (3.2) | 968 (3.8) | 190 (5.2) | 3133 (4.3) | 2275 (4.1) | 718 (4.5) | 140 (7.6) | 908 (1.9) | 608 (1.7) | 250 (2.6) | 50 (208) |
| **DD asthma** | 11208 (8.4) | 8351 (8.3) | 2480 (8.7) | 377 (9.2) | 7058 (8.7) | 5258 (8.5) | 1608 (9.1) | 192 (9.3) | 4150 (8.0) | 3093 (8.0) | 872 (8.1) | 185 (9.2) |
| **Occupational Sectors\*** |  |  |  |  |  |  |  |  |  |  |  |  |
| **Hair&beauty** |  |  | 1948 (6.8) | 58 (1.4) |  |  | 1888 (10.6) | 58 (2.8) |  |  | 60 (0.6) | 0 (0.0) |
| **Medical/dental** |  |  | 7898 (27.6) | 831 (20.3) |  |  | 7023 (39.5) | 687 (33.1) |  |  | 875 (8.1) | 144 (7.1) |
| **Laboratory** |  |  | 920 (3.2) | 2 (0.0) |  |  | 565 (3.2) | 1 (0.0) |  |  | 355 (3.3) | 1 (0.0) |
| **Textiles** |  |  | 82 (0.3) | 987 (24.1) |  |  | 33 (0.2) | 757 (26.5) |  |  | 49 (0.5) | 230 (11.4) |
| **Construction** |  |  | 4461 (15.6) | 1196 (29.2) |  |  | 37 (0.2) | 23 (1.1) |  |  | 4424 (40.7) | 1173 (57.9) |
| **Industry** |  |  | 2327 (8.1) | 993 (24.2) |  |  | 504 (2.8) | 564 (27.2) |  |  | 1823 (16.8) | 429 (21.2) |
| **Transport** |  |  | 3365 (11.7) | 46 (1.1) |  |  | 598 (3.4) | 5 (0.2) |  |  | 2767 (25.5) | 41 (2.0) |
| **Cleaning** |  |  | 7437 (26.0) | 191 (4.7) |  |  | 7017 (39.5) | 98 (4.7) |  |  | 420 (3.9) | 93 (4.6) |
| **Other** |  |  | 549 (1.9) | 8 (0.2) |  |  | 304 (1.7) | 3 (0.1) |  |  | 245 (2.3) | 5 (0.2) |

Mean (SD) for continuous variables, N (%) for categorical variables. \* Occupational sectors: The numbers do not add up to the total for each exposure group since some subjects have more than 1 job with MNP exposure. <sup>a</sup> Exposures were based on the OAsJEM [62]: organic dust=highest level of animals, flour, foods, plants-related dusts, moulds, endotoxin; gasses/fumes=highest level of exhaust fumes, high-level chemical disinfectants, bleach; pesticides=highest level of herbicides, insecticides, fungicides; metals=highest level of metal, metal working fluids; solvents=organic solvents. <sup>b</sup> Exposure is based on the EUROJEM respirable crystalline silica [63].

**Supplementary table S6.** Descriptives statistics stratified by MNP exposure for subjects included in the analysis on lung function decline – Total group and stratified by sex.

|  | Total group |  |  |  | Females |  |  |  | Males |  |  |  |
| --- | --- | --- | --- | --- | --- | --- | --- | --- | --- | --- | --- | --- |
|  | All | No exposure | Low exposure | High exposure | All Females | No exposure | Low exposure | High exposure | All Males | No exposure | Low exposure | High exposure |
| **N** | 45504 | 34561 (76.0) | 9647 (21.2) | 1296 (2.8) | 27837 | 21052 (75.6) | 6102 (21.9) | 683 (2.5) | 17667 | 13509 (76.5) | 3545 (20.1) | 613 (3.5) |
| **Age, yr** | 45.6 (11.0) | 45.4 (11.0) | 46.1 (11.0) | 46.2 (11.2) | 45.4 (10.7) | 45.1 (10.7) | 46.4 (10.8) | 46.4 (11.6) | 45.9 (11.4) | 45.9 (11.4) | 45.7 (11.3) | 46.1 (10.7) |
| **Male sex** | 17667 (38.8) | 13509 (76.5) | 3545 (20.1) | 613 (3.5) |  |  |  |  |  |  |  |  |
| **Height, cm** | 174.5 (9.3) | 174.8 (9.3) | 173.7 (9.1) | 174.5 (9.6) | 169.4 (6.4) | 169.6 (6.4) | 168.9 (6.4) | 168.2 (6.9) | 182.7 (7.0) | 182.9 (7.0) | 181.9 (7.0) | 181.6 (6.9) |
| **Weight, kg** | 79.1 (14.7) | 79.1 (14.6) | 79.2 (14.9) | 80.9 (15.4) | 73.7 (13.2) | 73.5 (13.1) | 74.0 (13.4) | 74.4 (14.6) | 87.8 (12.6) | 87.6 (12.4) | 88.1 (13.2) | 88.0 (12.9) |
| **BMI, kg/m<sup>2</sup>** | 25.9 (4.1) | 25.8 (4.0) | 26.2 (4.3) | 26.5 (4.4) | 25.7 (33.5) | 25.6 (4.4) | 26.0 (4.6) | 26.3 (5.1) | 26.3 (3.4) | 26.2 (3.4) | 26.6 (3.6) | 26.7 (3.5) |
| **Follow-up time, yr** | 8.5 (3.5) | 8.6 (3.5) | 8.5 (3.5) | 8.4 (3.6) | 8.5 (3.5) | 8.5 (3.5) | 8.3 (3.5) | 8.2 (3.5) | 8.6 (3.5) | 8.6 (3.5) | 8.6 (3.6) | 8.7 (3.6) |
| **Smoking** |  |  |  |  |  |  |  |  |  |  |  |  |
| **Never** | 20511 (45.1) | 15880 (45.9) | 4093 (42.4) | 538 (41.5) | 12837 (46.1) | 9829 (46.7) | 2700 (44.2) | 308 (45.1) | 7674 (43.4) | 6051 (44.8) | 1393 (39.3) | 230 (37.5) |
| **Ex** | 15745 (34.6) | 11790 (34.1) | 3521 (36.5) | 434 (33.5) | 9657 (34.7) | 7171 (34.1) | 2263 (37.1) | 223 (32.7) | 6088 (34.5) | 4619 (34.2) | 1258 (35.5) | 211 (34.4) |
| **Current** | 8314 (18.3) | 6166 (17.8) | 1846 (19.1) | 302 (23.3) | 4770 (17.1) | 3612 (17.2) | 1019 (16.7) | 139 (20.4) | 3544 (20.1) | 2554 (18.9) | 827 (23.3) | 163 (26.6) |
| **Missing** | 934 (2.1) | 725 (2.1) | 187 (1.9) | 22 (1.7) | 573 (2.1) | 440 (2.1) | 120 (2.0) | 13 (1.9) | 361 (2.0) | 285 (2.1) | 67 (1.9) | 9 (1.5) |
| **Education** |  |  |  |  |  |  |  |  |  |  |  |  |
| **Low** | 12781 (28.1) | 8064 (23.3) | 3964 (41.1) | 753 (58.1) | 7843 (28.2) | 5237 (24.9) | 2234 (36.6) | 372 (54.5) | 4938 (28.0) | 2827 (20.9) | 1730 (48.8) | 381 (62.2) |
| **Medium** | 17474 (38.4) | 13490 (39.0) | 3591 (37.2) | 393 (30.3) | 11041 (39.7) | 8523 (40.5) | 2290 (37.5) | 228 (33.4) | 6433 (36.4) | 4967 (36.8) | 1301 (36.7) | 165 (26.9) |
| **High** | 14545 (32.0) | 12506 (36.2) | 1923 (19.9) | 116 (9.0) | 8504 (30.5) | 6973 (33.1) | 1461 (23.9) | 70 (10.2) | 6041 (34.2) | 5533 (41.0) | 462 (13.0) | 46 (7.5) |
| **Missing** | 704 (1.5) | 501 (1.4) | 169 (1.8) | 34 (2.6) | 449 (1.6) | 319 (1.5) | 117 (1.9) | 13 (1.9) | 255 (1.4) | 182 (1.3) | 52 (1.5) | 21 (3.4) |
| **Income** |  |  |  |  |  |  |  |  |  |  |  |  |
| **Low** | 5031 (11.1) | 3551 (10.3) | 1257 (13.0) | 223 (17.2) | 3620 (13.0) | 2627 (12.5) | 850 (13.9) | 143 (20.9) | 1411 (8.0) | 924 (6.8) | 407 (11.5) | 80 (13.1) |
| **Medium** | 12374 (27.2) | 8744 (25.3) | 3218 (33.4) | 412 (31.8) | 7175 (25.8) | 5196 (24.7) | 1791 (29.4) | 188 (27.5) | 5199 (29.4) | 3548 (26.3) | 1427 (40.3) | 224 (36.5) |
| **High** | 20947 (46.0) | 17075 (49.4) | 3508 (36.4) | 364 (28.1) | 12220 (43.9) | 9767 (46.4) | 2268 (37.2) | 185 (27.1) | 8727 (49.4) | 7308 (54.1) | 1240 (35.0) | 179 (29.2) |
| **Unknown** | 7152 (15.7) | 5191 (15.0) | 1664 (17.2) | 297 (22.9) | 4822 (17.3) | 3462 (16.4) | 1193 (19.6) | 167 (24.5) | 2330 (13.2) | 1729 (12.8) | 471 (13.3) | 130 (21.2) |
| **Organic dust exposure<sup>a</sup>** |  |  |  |  |  |  |  |  |  |  |  |  |
| **No** | 42129 (92.6) | 32580 (94.3) | 8404 (87.1) | 1145 (88.3) | 27110 (97.4) | 20478 (97.3) | 5961 (97.7) | 671 (98.2) | 15019 (85.0) | 12102 (89.6) | 2443 (68.9) | 474 (77.3) |
| **Low** | 1591 (3.5) | 489 (1.4) | 977 (10.1) | 125 (9.6) | 197 (0.7) | 157 (0.7) | 32 (0.5) | 8 (1.2) | 1394 (7.9) | 332 (2.5) | 945 (26.7) | 117 (19.1) |
| **High** | 1784 (3.9) | 1492 (4.3) | 266 (2.8) | 26 (2.0) | 530 (1.9) | 417 (2.0) | 109 (1.8) | 4 (0.6) | 1254 (7.1) | 1075 (8.0) | 157 (4.4) | 22 (3.6) |
| **Gasses/Fumes exposure<sup>a</sup>** |  |  |  |  |  |  |  |  |  |  |  |  |
| **No** | 33252 (73.1) | 29365 (85.0) | 2986 (31.0) | 901 (69.5) | 19724 (70.9) | 18247 (86.7) | 1104 (18.1) | 373 (54.6) | 13528 (76.6) | 11118 (82.3) | 1882 (53.1) | 528 (86.1) |
| **Low** | 6729 (14.8) | 2130 (6.2) | 4241 (44.0) | 358 (27.6) | 4564 (16.4) | 754 (3.6) | 3517 (57.6) | 293 (42.9) | 2165 (12.3) | 1376 (10.2) | 724 (20.4) | 65 (10.6) |
| **High** | 5523 (12.1) | 3066 (8.9) | 2420 (25.1) | 37 (2.9) | 3549 (12.7) | 2051 (9.7) | 1481 (24.3) | 17 (2.5) | 1974 (11.2) | 1015 (7.5) | 939 (26.5) | 20 (3.3) |
| **Pesticide exposure<sup>a</sup>** |  |  |  |  |  |  |  |  |  |  |  |  |
| **No** | 43529 (95.7) | 32753 (94.8) | 9496 (98.4) | 1280 (98.8) | 27265 (97.9) | 20535 (97.5) | 6053 (99.2) | 677 (99.1) | 16264 (92.1) | 12218 (90.4) | 3443 (97.1) | 603 (98.4) |
| **Low** | 1187 (2.6) | 1082 (3.1) | 100 (1.0) | 5 (0.4) | 341 (1.2) | 310 (1.5) | 29 (0.5) | 2 (0.3) | 846 (4.8) | 772 (5.7) | 71 (2.0) | 3 (0.5) |
| **High** | 788 (1.7) | 726 (2.1) | 51 (0.5) | 11 (0.8) | 231 (0.8) | 207 (1.0) | 20 (0.3) | 4 (0.6) | 557 (3.2) | 519 (3.8) | 31 (0.9) | 7 (1.1) |
| **Metals exposure<sup>a</sup>** |  |  |  |  |  |  |  |  |  |  |  |  |
| **No** | 43638 (95.9) | 33709 (97.5) | 8719 (90.4) | 1210 (93.4) | 27682 (99.4) | 21034 (99.9) | 5987 (98.1) | 661 (96.8) | 15956 (90.3) | 12675 (93.8) | 2732 (77.1) | 549 (89.6) |
| **Low** | 1482 (3.3) | 617 (1.8) | 785 (8.1) | 80 (6.2) | 150 (0.5) | 16 (0.1) | 112 (1.8) | 22 (3.2) | 1332 (7.5) | 601 (4.4) | 673 (19.0) | 58 (9.5) |
| **High** | 384 (0.8) | 235 (0.7) | 143 (1.5) | 6 (0.5) | 5 (0.0) | 2 (0.0) | 3 (0.0) | 0 (0.0) | 379 (2.1) | 233 (1.7) | 140 (3.9) | 6 (1.0) |
| **Solvents exposure<sup>a</sup>** |  |  |  |  |  |  |  |  |  |  |  |  |
| **No** | 39349 (86.5) | 33079 (95.7) | 5475 (56.8) | 795 (61.3) | 24609 (88.4) | 20408 (96.9) | 3637 (59.6) | 564 (82.6) | 14740 (83.4) | 12671 (93.8) | 1838 (51.8) | 231 (37.7) |
| **Low** | 5047 (11.1) | 1350 (3.9) | 3499 (36.3) | 198 (15.3) | 2477 (8.9) | 601 (2.9) | 1820 (29.8) | 56 (8.2) | 2570 (14.5) | 749 (5.5) | 1679 (47.4) | 142 (23.2) |
| **High** | 1108 (2.4) | 132 (0.4) | 673 (7.0) | 303 (23.4) | 751 (2.7) | 43 (0.2) | 645 (10.6) | 63 (9.2) | 357 (2.0) | 89 (0.7) | 28 (0.8) | 240 (39.2) |
| **Silica exposure<sup>b</sup>** |  |  |  |  |  |  |  |  |  |  |  |  |
| **No** | 42654 (93.7) | 33933 (98.2) | 7746 (80.3) | 975 (75.2) | 27665 (99.4) | 20937 (99.5) | 6057 (99.3) | 671 (98.2) | 14989 (84.8) | 12996 (96.2) | 1689 (47.6) | 304 (49.6) |
| **Low** | 1128 (2.5) | 160 (0.5) | 809 (8.4) | 159 (12.3) | 61 (0.2) | 28 (0.1) | 26 (0.4) | 7 (1.0) | 1067 (6.0) | 132 (1.0) | 783 (22.1) | 152 (24.8) |
| **High** | 1722 (3.8) | 468 (1.4) | 1092 (11.3) | 162 (12.5) | 111 (0.4) | 87 (0.4) | 19 (0.3) | 5 (0.7) | 1611 (9.1) | 381 (2.8) | 1073 (30.3) | 157 (25.6) |
| **Lung function changes over time<sup>c</sup>** |  |  |  |  |  |  |  |  |  |  |  |  |
| **ΔFEV1, ml/yr** | -26.8 (39.1) | -26.6 (39.4) | -27.1 (37.7) | -28.0 (39.6) | -25.7 (33.5) | -25.5 (34.0) | -26.4 (31.8) | -26.7 (31.9) | -28.4 (46.5) | -28.4 (46.6) | -28.2 (46.0) | -29.4 (46.7) |
| **ΔFVC, ml/yr** | -23.6 (52.7) | -23.3 (53.2) | -24.5 (50.5) | -25.4 (53.0) | -23.5 (46.0) | -23.2 (46.7) | -24.7 (44.0) | -24.9 (43.9) | -23.7 (61.7) | -23.5 (62.1) | -24.1 (60.1) | -25.8 (61.6) |
| **ΔFEV1%FVC, %/yr** | -0.21 (0.64) | -0.21 (0.64) | -0.21 (0.65) | -0.21 (0.69) | -0.21 (0.64) | -0.21 (0.64) | -0.21 (0.66) | -0.22 (0.67) | -0.21 (0.63) | -0.21 (0.63) | -0.20 (0.62) | -0.20 (0.71) |
| **Occupational Sectors\*** |  |  |  |  |  |  |  |  |  |  |  |  |
| **Hair&beauty** |  |  | 667 (6.9) | 12 (0.9) |  |  | 643 (10.5) | 12 (1.8) |  |  | 24 (0.7) | 0 (0.0) |
| **Medical/dental** |  |  | 2888 (29.9) | 300 (23.1) |  |  | 2587 (42.4) | 244 (35.7) |  |  | 301 (8.5) | 56 (9.1) |
| **Laboratory** |  |  | 383 (4.0) | 0 (0.0) |  |  | 228 (3.7) | 0 (0.0) |  |  | 155 (4.4) | 0 (0.0) |
| **Textiles** |  |  | 22 (0.2) | 343 (26.5) |  |  | 11 (0.2) | 265 (38.8) |  |  | 11 (0.3) | 78 (12.7) |
| **Construction** |  |  | 1474 (15.3) | 359 (27.7) |  |  | 13 (0.2) | 8 (1.2) |  |  | 1461 (41.2) | 351 (57.3) |
| **Industry** |  |  | 778 (8.1) | 275 (21.2) |  |  | 165 (2.7) | 155 (22.7) |  |  | 613 (17.3) | 120 (19.6) |
| **Transport** |  |  | 1061 (11.0) | 7 (0.5) |  |  | 206 (3.4) | 0 (0.0) |  |  | 855 (24.1) | 7 (1.1) |
| **Cleaning** |  |  | 2310 (23.9) | 56 (4.3) |  |  | 2209 (36.2) | 34 (5.0) |  |  | 101 (2.8) | 22 (3.6) |
| **Other** |  |  | 196 (2.0) | 4 (0.3) |  |  | 114 (1.9) | 2 (0.3) |  |  | 82 (2.3) | 2 (0.3) |

Mean (SD) for continuous variables, N (%) for categorical variables. \* Occupational sectors: The numbers do not add up to the total for each exposure group since some subjects have more than 1 job with MNP exposure. <sup>a</sup> Exposures were based on the OAsJEM [62]: organic dust=highest level of animals, flour, foods, plants-related dusts, moulds, endotoxin; gasses/fumes=highest level of exhaust fumes, high-level chemical disinfectants, bleach; pesticides=highest level of herbicides, insecticides, fungicides; metals=highest level of metal, metal working fluids; solvents=organic solvents. <sup>b</sup> Exposure is based on the EUROJEM respirable crystalline silica [63]. <sup>c</sup> calculated as (lung function at the most recent visit-lung function at baseline)/time between baseline and most recent visit.

**Supplementary table S7.** Development of airway obstruction, respiratory symptoms and asthma stratified by MNP exposure – Total group and stratified by sex.

|  | Total group |  |  |  | Females |  |  |  | Males |  |  |  |
| --- | --- | --- | --- | --- | --- | --- | --- | --- | --- | --- | --- | --- |
|  | All | No exposure | Low exposure | High exposure | All Females | No exposure | Low exposure | High exposure | All Males | No exposure | Low exposure | High exposure |
|  | n/N (%) | n/N (%) | n/N (%) | n/N (%) | n/N (%) | n/N (%) | n/N (%) | n/N (%) | n/N (%) | n/N (%) | n/N (%) | n/N (%) |
| **Airway obstruction** |  |  |  |  |  |  |  |  |  |  |  |  |
| **FEV1/FVC < 0.7** | 3632/40888<br>(8.9) | 2709/31268<br>(8.7) | 816/8514<br>(9.6) | 107/1106<br>(9.7) | 2127/25610<br>(8.3) | 1574/19517<br>(8.1) | 503/5505<br>(9.1) | 50/588<br>(8.5) | 1505/15278<br>(9.9) | 1135/11751<br>(9.7) | 313/3009<br>(10.4) | 57/518<br>(11.0) |
| **FEV1/FVC < LLN** | 1780/42168<br>(4.2) | 1325/32184<br>(4.1) | 409/8832<br>(4.6) | 46/1152<br>(4.0) | 1134/25963<br>(4.4) | 826/19715<br>(4.2) | 280/5644<br>(5.0) | 28/604<br>(4.6) | 646/16205<br>(4.0) | 499/12469<br>(4.0) | 129/3188<br>(4.0) | 18/548<br>(3.3) |
| **Respiratory symptoms** |  |  |  |  |  |  |  |  |  |  |  |  |
| **Wheeze** | 1949/62312<br>(3.1) | 1464/47299<br>(3.1) | 415/13227<br>(3.1) | 70/1768<br>(4.0) | 1139/38702<br>(2.9) | 859/29165<br>(2.9) | 251/8590<br>(2.9) | 29/947<br>(3.1) | 810/23610<br>(3.4) | 605/18134<br>(3.3) | 164/4637<br>(3.5) | 41/839<br>(4.9) |
| **Cough** | 4225/65630<br>(6.4) | 3126/49716<br>(6.3) | 942/14031<br>(6.7) | 157/1863<br>(8.3) | 2326/40672<br>(5.7) | 1740/30618<br>(5.7) | 511/9052<br>(5.6) | 75/1002<br>(7.5) | 1899/24958<br>(7.6) | 1386/19098<br>(7.3) | 431/4979<br>(8.7) | 82/881<br>(9.3) |
| **Sputum** | 3384/66424<br>(5.1) | 2515/50337<br>(5.0) | 758/14165<br>(5.4) | 111/1922<br>(5.8) | 1817/41367<br>(4.4) | 1373/31150<br>(4.4) | 401/9198<br>(4.4) | 43/1019<br>(4.2) | 1567/25057<br>(6.3) | 1142/19187<br>(6.0) | 357/4967<br>(7.2) | 68/903<br>(7.5) |
| **Bronchitis** | 2355/68688<br>(3.4) | 1713/52008<br>(3.3) | 562/14673<br>(3.8) | 80/2007<br>(4.0) | 1285/42541<br>(3.0) | 953/32021<br>(3.0) | 303/9467<br>(3.2) | 29/1053<br>(2.8) | 1070/26147<br>(4.1) | 760/19987<br>(3.8) | 259/5206<br>(5.0) | 51/954<br>(5.3) |
| **DD asthma** | 2494/65118<br>(3.8) | 1820/49343<br>(3.7) | 588/13870<br>(4.2) | 86/1905<br>(4.5) | 1581/40172<br>(3.9) | 1151/30286<br>(3.8) | 376/8888<br>(4.2) | 54/998<br>(5.4) | 913/24946<br>(3.7) | 669/19057<br>(3.5) | 212/4982<br>(4.3) | 32/907<br>(3.5) |

n/N: number of cases/total number of subjects in analysis

**Supplementary table S8.** Associations between occupational MNP exposure and lung function and airway obstruction – Base analysis: no adjustment for co-exposures and no adjustment for education and income - Total group and stratified by sex.

|  | Total group |  |  |  | Females |  |  |  | Males |  |  |  |
| --- | --- | --- | --- | --- | --- | --- | --- | --- | --- | --- | --- | --- |
|  | n | estimate | 95% CI | P | n | estimate | 95% CI | P | n | estimate | 95% CI | P |
| **Outcome: FEV1 (ml)** |  |  |  |  |  |  |  |  |  |  |  |  |
| No exposure | 71358 | reference |  |  | 43951 | reference |  |  | 27407 | reference |  |  |
| Low exposure | 20453 | -14.691 | -21.908;-7.475 | <0.001 | 12748 | -10.966 | -18.642;-3.289 | 0.005 | 7705 | -27.069 | -41.114;-13.023 | <0.001 |
| High exposure | 3033 | -51.623 | -68.491;-34.754 | <0.001 | 1576 | -53.978 | -73.518;-34.438 | <0.001 | 1457 | -49.777 | -79.022;-20.532 | <0.001 |
| **Outcome: FVC (ml)** |  |  |  |  |  |  |  |  |  |  |  |  |
| No exposure | 71358 | reference |  |  | 43951 | reference |  |  | 27407 | reference |  |  |
| Low exposure | 20453 | -6.453 | -14.859;1.952 | 0.132 | 12748 | -5.503 | -14.603;3.597 | 0.236 | 7705 | -13.300 | -29.417;2.817 | 0.106 |
| High exposure | 3033 | -49.688 | -69.336;-30.040 | <0.001 | 1576 | -59.146 | -82.310;-35.982 | <0.001 | 1457 | -36.941 | -70.499;-3.383 | 0.031 |
| **Outcome: FEV1%FVC (%)** |  |  |  |  |  |  |  |  |  |  |  |  |
| No exposure | 71358 | reference |  |  | 43951 | reference |  |  | 27407 | reference |  |  |
| Low exposure | 20453 | -0.238 | -0.338;-0.138 | <0.001 | 12748 | -0.188 | -0.310;-0.066 | 0.003 | 7705 | -0.312 | -0.484;-0.140 | <0.001 |
| High exposure | 3033 | -0.328 | -0.562;-0.094 | 0.006 | 1576 | -0.294 | -0.606;0.017 | 0.064 | 1457 | -0.364 | -0.722;-0.005 | 0.047 |
|  |  | OR | 95% CI | P |  | OR | 95% CI | P |  | OR | 95% CI | P |
| **Outcome: FEV1/FVC<0.7** |  |  |  |  |  |  |  |  |  |  |  |  |
| No exposure | 71358 | reference |  |  | 43951 | reference |  |  | 27407 | reference |  |  |
| Low exposure | 20453 | 1.079 | 1.031;1.129 | 0.001 | 12748 | 1.064 | 1.002;1.131 | 0.044 | 7705 | 1.104 | 1.030;1.182 | 0.005 |
| High exposure | 3033 | 1.157 | 1.046;1.280 | 0.005 | 1576 | 1.247 | 1.079;1.440 | 0.003 | 1457 | 1.084 | 0.940;1.249 | 0.266 |
| **Outcome: FEV1/FVC<LLN** |  |  |  |  |  |  |  |  |  |  |  |  |
| No exposure | 71358 | reference |  |  | 43951 | reference |  |  | 27407 | reference |  |  |
| Low exposure | 20453 | 1.062 | 1.012;1.114 | 0.014 | 12748 | 1.033 | 0.970;1.099 | 0.312 | 7705 | 1.108 | 1.028;1.195 | 0.008 |
| High exposure | 3033 | 1.151 | 1.034;1.280 | 0.010 | 1576 | 1.205 | 1.040;1.397 | 0.013 | 1457 | 1.096 | 0.939;1.280 | 0.244 |

Linear regression models with lung function as outcome, logistic regression models with airway obstruction as outcome. All models are adjusted for age, sex, height, and smoking.

**Supplementary table S9.** Associations between occupational MNP exposure and lung function and airway obstruction – Main analysis - Total group and stratified by sex.

|  | Total group |  |  |  | Females |  |  |  | Males |  |  |  |
| --- | --- | --- | --- | --- | --- | --- | --- | --- | --- | --- | --- | --- |
|  | n | estimate | 95% CI | P | n | estimate | 95% CI | P | n | estimate | 95% CI | P |
| **Outcome: FEV1 (ml)** |  |  |  |  |  |  |  |  |  |  |  |  |
| No exposure | 71358 | reference |  |  | 43951 | reference |  |  | 27407 | reference |  |  |
| Low exposure | 20453 | -5.686 | -16.332;4.961 | 0.295 | 12748 | -17.839 | -30.023;-5.655 | 0.004 | 7705 | -17.414 | -38.156;3.328 | 0.100 |
| High exposure | 3033 | -42.920 | -61.248;-24.593 | <0.001 | 1576 | -63.724 | -84.155;-43.293 | <0.001 | 1457 | -43.875 | -79.355;-8.395 | 0.015 |
| **Outcome: FVC (ml)** |  |  |  |  |  |  |  |  |  |  |  |  |
| No exposure | 71358 | reference |  |  | 43951 | reference |  |  | 27407 | reference |  |  |
| Low exposure | 20453 | -3.706 | -16.110;8.698 | 0.558 | 12748 | -20.555 | -34.999;-6.112 | 0.005 | 7705 | -9.878 | -33.686;13.930 | 0.416 |
| High exposure | 3033 | -47.415 | -68.768;-26.063 | <0.001 | 1576 | -73.197 | -97.416;-48.978 | <0.001 | 1457 | -41.323 | -82.047;-0.599 | 0.047 |
| **Outcome: FEV1%FVC (%)** |  |  |  |  |  |  |  |  |  |  |  |  |
| No exposure | 71358 | reference |  |  | 43951 | reference |  |  | 27407 | reference |  |  |
| Low exposure | 20453 | -0.140 | -0.288;0.007 | 0.063 | 12748 | -0.132 | -0.326;0.062 | 0.182 | 7705 | -0.160 | -0.414;0.095 | 0.219 |
| High exposure | 3033 | -0.256 | -0.511;-0.002 | 0.049 | 1576 | -0.304 | -0.630;0.022 | 0.067 | 1457 | -0.216 | -0.652;0.219 | 0.331 |
|  |  | OR | 95% CI | P |  | OR | 95% CI | P |  | OR | 95% CI | P |
| **Outcome: FEV1/FVC<0.7** |  |  |  |  |  |  |  |  |  |  |  |  |
| No exposure | 71358 | reference |  |  | 43951 | reference |  |  | 27407 | reference |  |  |
| Low exposure | 20453 | 1.092 | 1.022;1.166 | 0.010 | 12748 | 1.086 | 0.988;1.194 | 0.089 | 7705 | 1.130 | 1.021;1.251 | 0.018 |
| High exposure | 3033 | 1.184 | 1.060;1.322 | 0.003 | 1576 | 1.276 | 1.100;1.482 | 0.001 | 1457 | 1.100 | 0.925;1.309 | 0.281 |
| **Outcome: FEV1/FVC<LLN** |  |  |  |  |  |  |  |  |  |  |  |  |
| No exposure | 71358 | reference |  |  | 43951 | reference |  |  | 27407 | reference |  |  |
| Low exposure | 20453 | 1.055 | 0.984;1.132 | 0.134 | 12748 | 1.052 | 0.955;1.160 | 0.304 | 7705 | 1.104 | 0.988;1.235 | 0.081 |
| High exposure | 3033 | 1.155 | 1.028;1.298 | 0.015 | 1576 | 1.232 | 1.056;1.437 | 0.008 | 1457 | 1.058 | 0.874;1.281 | 0.563 |

Linear regression models with lung function as outcome, logistic regression models with airway obstruction as outcome. All models are adjusted for age, sex, height, smoking, and occupational exposure to organic dust, gasses/fumes, pesticides, metals, solvents and silica.

**Supplementary table S10.**
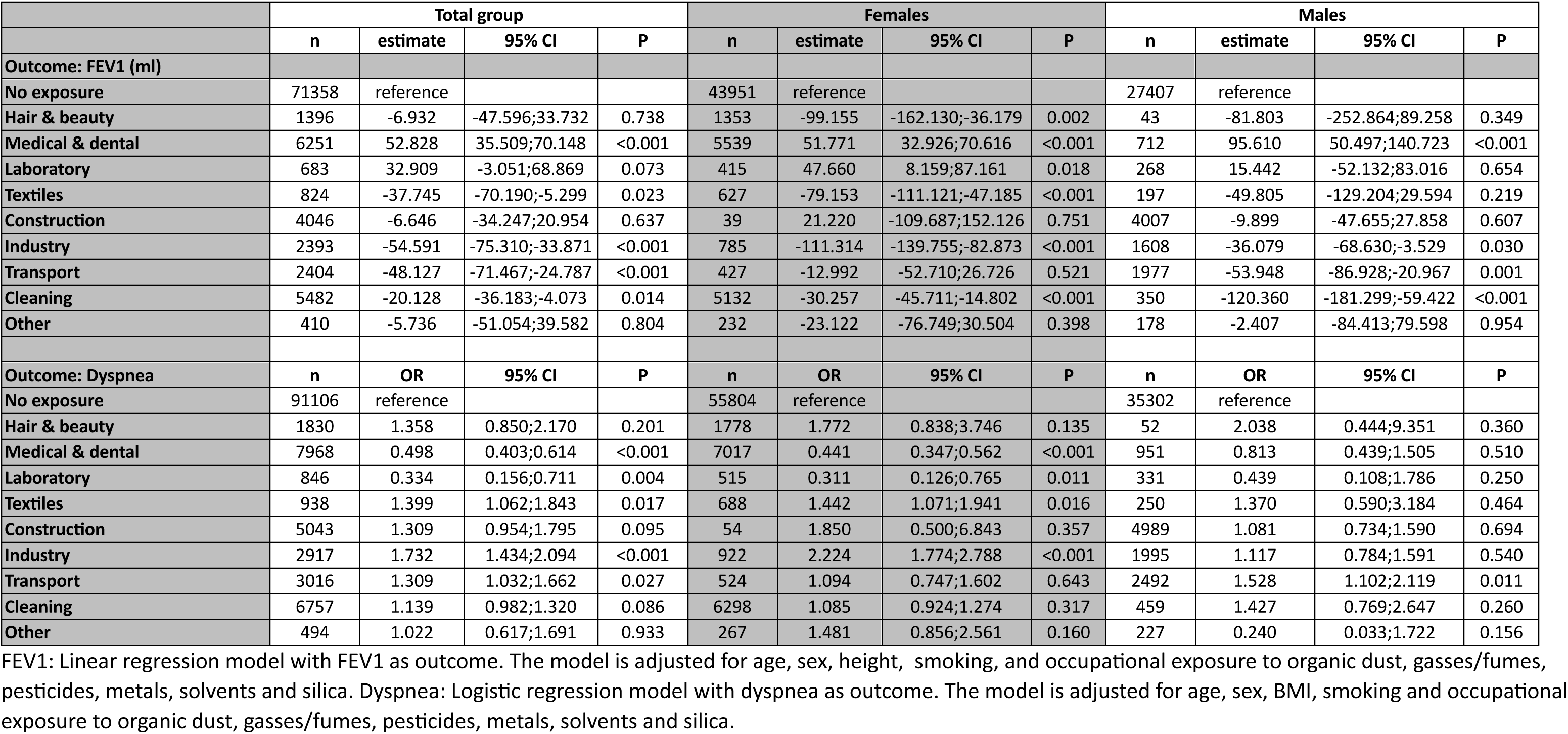
Associations between occupational MNP exposure in specific occupational sectors and FEV1/Dyspnea – Main analysis - Total group and stratified by sex.

**Supplementary table S11.**
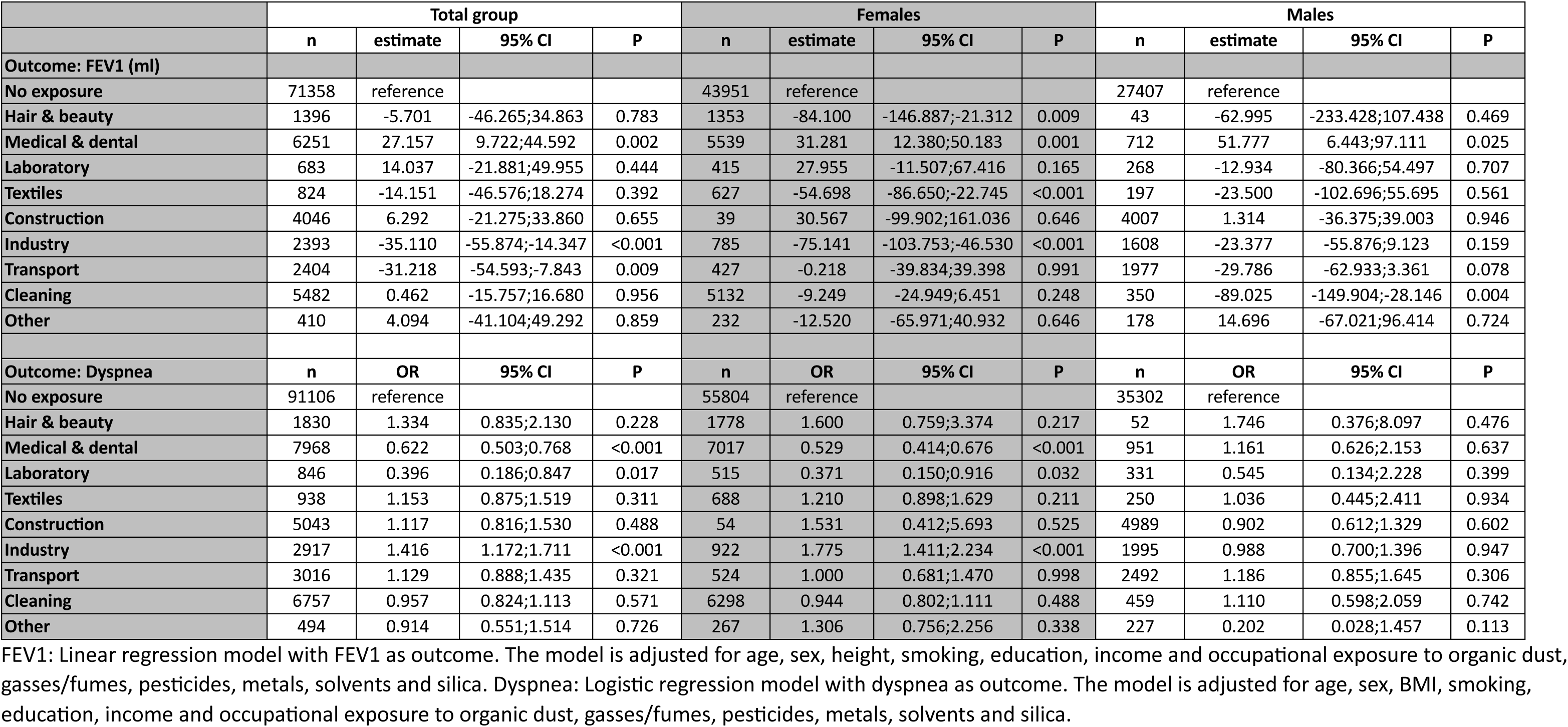
Associations between occupational MNP exposure in specific occupational sectors and FEV1/Dyspnea – Sensitivity analysis: additional adjustment for education and income - Total group and stratified by sex.

**Supplementary table S12.** Associations between occupational MNP exposure and lung function and airway obstruction – Sensitivity analysis: additional adjustment for education and income - Total group and stratified by sex.

|  | Total group |  |  |  | Females |  |  |  | Males |  |  |  |
| --- | --- | --- | --- | --- | --- | --- | --- | --- | --- | --- | --- | --- |
|  | n | estimate | 95% CI | P | n | estimate | 95% CI | P | n | estimate | 95% CI | P |
| **Outcome: FEV1 (ml)** |  |  |  |  |  |  |  |  |  |  |  |  |
| No exposure | 71358 | reference |  |  | 43951 | reference |  |  | 27407 | reference |  |  |
| Low exposure | 20453 | -0.509 | -11.160;10.143 | 0.925 | 12748 | -8.630 | -20.859;3.599 | 0.167 | 7705 | -10.728 | -31.443;9.987 | 0.310 |
| High exposure | 3033 | -21.505 | -39.888;-3.123 | 0.022 | 1576 | -36.161 | -56.671;-15.651 | <0.001 | 1457 | -22.866 | -58.360;12.628 | 0.207 |
| **Outcome: FVC (ml)** |  |  |  |  |  |  |  |  |  |  |  |  |
| No exposure | 71358 | reference |  |  | 43951 | reference |  |  | 27407 | reference |  |  |
| Low exposure | 20453 | 3.723 | -8.668;16.114 | 0.556 | 12748 | -6.964 | -21.434;7.505 | 0.346 | 7705 | -1.843 | -25.595;21.908 | 0.879 |
| High exposure | 3033 | -19.101 | -40.486;2.283 | 0.080 | 1576 | -36.681 | -60.950;-12.413 | 0.003 | 1457 | -16.136 | -56.833;24.562 | 0.437 |
| **Outcome: FEV1%FVC (%)** |  |  |  |  |  |  |  |  |  |  |  |  |
| No exposure | 71358 | reference |  |  | 43951 | reference |  |  | 27407 | reference |  |  |
| Low exposure | 20453 | -0.139 | -0.287;0.009 | 0.066 | 12748 | -0.144 | -0.339;0.052 | 0.150 | 7705 | -0.147 | -0.402;0.109 | 0.260 |
| High exposure | 3033 | -0.223 | -0.479;0.033 | 0.087 | 1576 | -0.287 | -0.616;0.041 | 0.086 | 1457 | -0.172 | -0.609;0.266 | 0.442 |
|  |  | OR | 95% CI | P |  | OR | 95% CI | P |  | OR | 95% CI | P |
| **Outcome: FEV1/FVC<0.7** |  |  |  |  |  |  |  |  |  |  |  |  |
| No exposure | 71358 | reference |  |  | 43951 | reference |  |  | 27407 | reference |  |  |
| Low exposure | 20453 | 1.081 | 1.012;1.155 | 0.021 | 12748 | 1.079 | 0.981;1.187 | 0.119 | 7705 | 1.114 | 1.006;1.233 | 0.037 |
| High exposure | 3033 | 1.141 | 1.021;1.276 | 0.020 | 1576 | 1.231 | 1.059;1.431 | 0.007 | 1457 | 1.067 | 0.896;1.270 | 0.469 |
| **Outcome: FEV1/FVC<LLN** |  |  |  |  |  |  |  |  |  |  |  |  |
| No exposure | 71358 | reference |  |  | 43951 | reference |  |  | 27407 | reference |  |  |
| Low exposure | 20453 | 1.042 | 0.971;1.118 | 0.250 | 12748 | 1.040 | 0.943;1.147 | 0.430 | 7705 | 1.090 | 0.974;1.218 | 0.133 |
| High exposure | 3033 | 1.104 | 0.982;1.242 | 0.097 | 1576 | 1.174 | 1.005;1.371 | 0.043 | 1457 | 1.020 | 0.842;1.236 | 0.836 |

Linear regression models with lung function as outcome, logistic regression models with airway obstruction as outcome. All models are adjusted for age, sex, height, education, income, smoking and occupational exposure to organic dust, gasses/fumes, pesticides, metals, solvents and silica.

**Supplementary table S13.** Associations between occupational MNP exposure and respiratory symptoms and asthma – Base analysis: no adjustment for co-exposures and no adjustment for education and income - Total group and stratified by sex.

|  | Total group |  |  |  | Females |  |  |  | Males |  |  |  |
| --- | --- | --- | --- | --- | --- | --- | --- | --- | --- | --- | --- | --- |
|  | n | OR | 95% CI | P | n | OR | 95% CI | P | n | OR | 95% CI | P |
| **Outcome: Wheeze** |  |  |  |  |  |  |  |  |  |  |  |  |
| No exposure | 100232 | reference |  |  | 61506 | reference |  |  | 38726 | reference |  |  |
| Low exposure | 28491 | 1.068 | 1.026;1.111 | 0.001 | 17699 | 1.041 | 0.989;1.096 | 0.125 | 10792 | 1.107 | 1.040;1.178 | 0.001 |
| High exposure | 4058 | 1.152 | 1.053;1.260 | 0.002 | 2060 | 1.076 | 0.944;1.226 | 0.274 | 1998 | 1.231 | 1.087;1.393 | 0.001 |
| **Outcome: Cough** |  |  |  |  |  |  |  |  |  |  |  |  |
| No exposure | 100054 | reference |  |  | 61412 | reference |  |  | 38642 | reference |  |  |
| Low exposure | 28456 | 1.016 | 0.967;1.068 | 0.529 | 17656 | 0.990 | 0.927;1.057 | 0.756 | 10800 | 1.048 | 0.970;1.133 | 0.233 |
| High exposure | 4068 | 1.251 | 1.125;1.391 | <0.001 | 2063 | 1.195 | 1.024;1.393 | 0.023 | 2005 | 1.294 | 1.118;1.499 | <0.001 |
| **Outcome: Sputum** |  |  |  |  |  |  |  |  |  |  |  |  |
| No exposure | 100092 | reference |  |  | 61446 | reference |  |  | 38646 | reference |  |  |
| Low exposure | 28482 | 1.074 | 1.018;1.134 | 0.009 | 17697 | 1.050 | 0.976;1.130 | 0.191 | 10785 | 1.103 | 1.019;1.194 | 0.016 |
| High exposure | 4070 | 1.170 | 1.040;1.317 | 0.009 | 2061 | 1.081 | 0.901;1.299 | 0.402 | 2009 | 1.232 | 1.055;1.439 | 0.008 |
| **Outcome: Bronchitis** |  |  |  |  |  |  |  |  |  |  |  |  |
| No exposure | 100453 | reference |  |  | 61647 | reference |  |  | 38806 | reference |  |  |
| Low exposure | 28584 | 1.040 | 0.966;1.118 | 0.298 | 17747 | 1.016 | 0.922;1.120 | 0.748 | 10837 | 1.068 | 0.956;1.192 | 0.246 |
| High exposure | 4097 | 1.250 | 1.073;1.457 | 0.004 | 2072 | 1.247 | 0.998;1.559 | 0.053 | 2025 | 1.239 | 1.004;1.529 | 0.045 |
| **Outcome: Dyspnea** |  |  |  |  |  |  |  |  |  |  |  |  |
| No exposure | 91106 | reference |  |  | 55804 | reference |  |  | 35302 | reference |  |  |
| Low exposure | 25702 | 1.069 | 0.990;1.153 | 0.088 | 15956 | 0.996 | 0.912;1.088 | 0.925 | 9746 | 1.334 | 1.146;1.553 | <0.001 |
| High exposure | 3630 | 1.568 | 1.341;1.835 | <0.001 | 1841 | 1.603 | 1.332;1.929 | <0.001 | 1789 | 1.528 | 1.136;2.055 | 0.005 |
| **Outcome: DD Asthma** |  |  |  |  |  |  |  |  |  |  |  |  |
| No exposure | 100321 | reference |  |  | 61566 | reference |  |  | 38755 | reference |  |  |
| Low exposure | 28531 | 1.057 | 1.008;1.108 | 0.021 | 17710 | 1.094 | 1.031;1.160 | 0.003 | 10821 | 1.003 | 0.927;1.085 | 0.943 |
| High exposure | 4087 | 1.129 | 1.012;1.259 | 0.029 | 2067 | 1.096 | 0.941;1.276 | 0.239 | 2020 | 1.166 | 0.997;1.364 | 0.054 |

Logistic regression models adjusted for age, sex, BMI and smoking.

**Supplementary table S14.** Associations between occupational MNP exposure and respiratory symptoms and asthma – Main analysis - Total group and stratified by sex.

|  | Total group |  |  |  | Females |  |  |  | Males |  |  |  |
| --- | --- | --- | --- | --- | --- | --- | --- | --- | --- | --- | --- | --- |
|  | n | OR | 95% CI | P | n | OR | 95% CI | P | n | OR | 95% CI | P |
| **Outcome: Wheeze** |  |  |  |  |  |  |  |  |  |  |  |  |
| No exposure | 100232 | reference |  |  | 61506 | reference |  |  | 38726 | reference |  |  |
| Low exposure | 28491 | 1.079 | 1.018;1.144 | 0.011 | 17699 | 1.057 | 0.974;1.146 | 0.183 | 10792 | 1.125 | 1.026;1.233 | 0.012 |
| High exposure | 4058 | 1.155 | 1.047;1.274 | 0.004 | 2060 | 1.079 | 0.941;1.236 | 0.276 | 1998 | 1.264 | 1.087;1.468 | 0.002 |
| **Outcome: Cough** |  |  |  |  |  |  |  |  |  |  |  |  |
| No exposure | 100054 | reference |  |  | 61412 | reference |  |  | 38642 | reference |  |  |
| Low exposure | 28456 | 0.968 | 0.900;1.040 | 0.372 | 17656 | 1.016 | 0.918;1.123 | 0.762 | 10800 | 0.920 | 0.821;1.032 | 0.157 |
| High exposure | 4068 | 1.218 | 1.084;1.369 | <0.001 | 2063 | 1.219 | 1.038;1.431 | 0.016 | 2005 | 1.139 | 0.950;1.365 | 0.159 |
| **Outcome: Sputum** |  |  |  |  |  |  |  |  |  |  |  |  |
| No exposure | 100092 | reference |  |  | 61446 | reference |  |  | 38646 | reference |  |  |
| Low exposure | 28482 | 0.988 | 0.914;1.069 | 0.772 | 17697 | 1.104 | 0.985;1.238 | 0.090 | 10785 | 0.911 | 0.809;1.025 | 0.120 |
| High exposure | 4070 | 1.087 | 0.954;1.239 | 0.212 | 2061 | 1.131 | 0.935;1.367 | 0.204 | 2009 | 0.995 | 0.821;1.206 | 0.960 |
| **Outcome: Bronchitis** |  |  |  |  |  |  |  |  |  |  |  |  |
| No exposure | 100453 | reference |  |  | 61647 | reference |  |  | 38806 | reference |  |  |
| Low exposure | 28584 | 0.966 | 0.869;1.074 | 0.521 | 17747 | 1.110 | 0.954;1.290 | 0.176 | 10837 | 0.854 | 0.724;1.007 | 0.060 |
| High exposure | 4097 | 1.186 | 1.001;1.404 | 0.049 | 2072 | 1.320 | 1.047;1.663 | 0.019 | 2025 | 0.938 | 0.720;1.223 | 0.638 |
| **Outcome: Dyspnea** |  |  |  |  |  |  |  |  |  |  |  |  |
| No exposure | 91106 | reference |  |  | 55804 | reference |  |  | 35302 | reference |  |  |
| Low exposure | 25702 | 1.045 | 0.935;1.168 | 0.440 | 15956 | 1.011 | 0.885;1.155 | 0.869 | 9746 | 1.160 | 0.924;1.457 | 0.202 |
| High exposure | 3630 | 1.582 | 1.337;1.871 | <0.001 | 1841 | 1.683 | 1.388;2.040 | <0.001 | 1789 | 1.499 | 1.044;2.151 | 0.028 |
| **Outcome: DD Asthma** |  |  |  |  |  |  |  |  |  |  |  |  |
| No exposure | 100321 | reference |  |  | 61566 | reference |  |  | 38755 | reference |  |  |
| Low exposure | 28531 | 1.042 | 0.972;1.118 | 0.242 | 17710 | 1.065 | 0.969;1.170 | 0.193 | 10821 | 1.047 | 0.934;1.173 | 0.431 |
| High exposure | 4087 | 1.152 | 1.023;1.296 | 0.019 | 2067 | 1.081 | 0.921;1.268 | 0.342 | 2020 | 1.191 | 0.987;1.438 | 0.069 |

Logistic regression models adjusted for age, sex, BMI, smoking and occupational exposure to organic dust, gasses/fumes, pesticides, metals, solvents and silica.

**Supplementary table S15.** Associations between occupational MNP exposure and respiratory symptoms and asthma – Sensitivity analysis: additional adjustment for education and income - Total group and stratified by sex.

|  | Total group |  |  |  | Females |  |  |  | Males |  |  |  |
| --- | --- | --- | --- | --- | --- | --- | --- | --- | --- | --- | --- | --- |
|  | n | OR | 95% CI | P | n | OR | 95% CI | P | n | OR | 95% CI | P |
| **Outcome: Wheeze** |  |  |  |  |  |  |  |  |  |  |  |  |
| No exposure | 100232 | reference |  |  | 61506 | reference |  |  | 38726 | reference |  |  |
| Low exposure | 28491 | 1.060 | 1.000;1.123 | 0.050 | 17699 | 1.031 | 0.950;1.118 | 0.471 | 10792 | 1.099 | 1.002;1.204 | 0.044 |
| High exposure | 4058 | 1.092 | 0.990;1.205 | 0.079 | 2060 | 1.014 | 0.884;1.164 | 0.837 | 1998 | 1.200 | 1.032;1.395 | 0.018 |
| **Outcome: Cough** |  |  |  |  |  |  |  |  |  |  |  |  |
| No exposure | 100054 | reference |  |  | 61412 | reference |  |  | 38642 | reference |  |  |
| Low exposure | 28456 | 0.932 | 0.867;1.002 | 0.057 | 17656 | 0.963 | 0.870;1.065 | 0.464 | 10800 | 0.889 | 0.792;0.997 | 0.044 |
| High exposure | 4068 | 1.099 | 0.978;1.235 | 0.114 | 2063 | 1.068 | 0.909;1.255 | 0.421 | 2005 | 1.059 | 0.883;1.270 | 0.539 |
| **Outcome: Sputum** |  |  |  |  |  |  |  |  |  |  |  |  |
| No exposure | 100092 | reference |  |  | 61446 | reference |  |  | 38646 | reference |  |  |
| Low exposure | 28482 | 0.963 | 0.891;1.041 | 0.345 | 17697 | 1.060 | 0.945;1.189 | 0.320 | 10785 | 0.886 | 0.787;0.996 | 0.043 |
| High exposure | 4070 | 1.001 | 0.877;1.141 | 0.991 | 2061 | 1.020 | 0.843;1.234 | 0.841 | 2009 | 0.936 | 0.772;1.135 | 0.499 |
| **Outcome: Bronchitis** |  |  |  |  |  |  |  |  |  |  |  |  |
| No exposure | 100453 | reference |  |  | 61647 | reference |  |  | 38806 | reference |  |  |
| Low exposure | 28584 | 0.926 | 0.834;1.029 | 0.155 | 17747 | 1.051 | 0.903;1.222 | 0.521 | 10837 | 0.818 | 0.694;0.964 | 0.016 |
| High exposure | 4097 | 1.062 | 0.896;1.258 | 0.491 | 2072 | 1.158 | 0.917;1.460 | 0.218 | 2025 | 0.860 | 0.659;1.121 | 0.264 |
| **Outcome: Dyspnea** |  |  |  |  |  |  |  |  |  |  |  |  |
| No exposure | 91106 | reference |  |  | 55804 | reference |  |  | 35302 | reference |  |  |
| Low exposure | 25702 | 0.961 | 0.860;1.073 | 0.479 | 15956 | 0.928 | 0.812;1.061 | 0.277 | 9746 | 1.016 | 0.811;1.273 | 0.891 |
| High exposure | 3630 | 1.272 | 1.074;1.506 | 0.005 | 1841 | 1.361 | 1.120;1.652 | 0.002 | 1789 | 1.165 | 0.814;1.669 | 0.404 |
| **Outcome: DD Asthma** |  |  |  |  |  |  |  |  |  |  |  |  |
| No exposure | 100321 | reference |  |  | 61566 | reference |  |  | 38755 | reference |  |  |
| Low exposure | 28531 | 1.029 | 0.960;1.104 | 0.415 | 17710 | 1.040 | 0.946;1.143 | 0.418 | 10821 | 1.035 | 0.923;1.160 | 0.554 |
| High exposure | 4087 | 1.107 | 0.984;1.247 | 0.091 | 2067 | 1.028 | 0.876;1.207 | 0.733 | 2020 | 1.160 | 0.960;1.401 | 0.124 |

Logistic regression models adjusted for age, sex, BMI, education, income, smoking and occupational exposure to organic dust, gasses/fumes, pesticides, metals, solvents and silica.

**Supplementary table S16.** Associations between occupational MNP exposure and lung function decline – Base analysis: no adjustment for co-exposures and no adjustment for education and income - Total group and stratified by sex. Restricted to subjects with at least two lung function measurements and ≥ 25 years at baseline.

|  | Total group |  |  |  | Females |  |  |  | Males |  |  |  |
| --- | --- | --- | --- | --- | --- | --- | --- | --- | --- | --- | --- | --- |
|  | n | estimate | 95% CI | P | n | estimate | 95% CI | P | n | estimate | 95% CI | P |
| **Outcome: $\Delta$ FEV1 (ml/yr)** |  |  |  |  |  |  |  |  |  |  |  |  |
| No exposure | 34561 | reference |  |  | 21052 | reference |  |  | 13509 | reference |  |  |
| Low exposure | 9647 | -0.687 | -1.304;-0.070 | 0.029 | 6102 | -0.654 | -1.335;0.027 | 0.060 | 3545 | -0.833 | -2.007;0.342 | 0.165 |
| High exposure | 1296 | -0.543 | -2.063;0.978 | 0.484 | 683 | -0.791 | -2.642;1.059 | 0.402 | 613 | -0.284 | -2.837;2.268 | 0.827 |
| **Outcome: $\Delta$ FVC (ml/yr)** |  |  |  |  |  |  |  |  |  |  |  |  |
| No exposure | 34561 | reference |  |  | 21052 | reference |  |  | 13509 | reference |  |  |
| Low exposure | 9647 | -1.243 | -2.042;-0.443 | 0.002 | 6102 | -0.975 | -1.866;-0.085 | 0.032 | 3545 | -1.688 | -3.202;-0.174 | 0.029 |
| High exposure | 1296 | -1.244 | -3.216;0.727 | 0.216 | 683 | -1.106 | -3.526;1.314 | 0.370 | 613 | -1.400 | -4.690;1.890 | 0.404 |
| **Outcome: $\Delta$ FEV1%FVC (%/yr)** |  |  |  |  |  |  |  |  |  |  |  |  |
| No exposure | 34561 | reference |  |  | 21052 | reference |  |  | 13509 | reference |  |  |
| Low exposure | 9647 | 0.003 | -0.007;0.012 | 0.589 | 6102 | 0.000 | -0.013;0.012 | 0.938 | 3545 | 0.004 | -0.011;0.019 | 0.618 |
| High exposure | 1296 | 0.004 | -0.019;0.028 | 0.709 | 683 | -0.004 | -0.037;0.030 | 0.830 | 613 | 0.013 | -0.020;0.046 | 0.449 |

Linear mixed regression models with lung function as outcome. All models are adjusted for age, sex, height and smoking at baseline. Random intercept. The presented estimates are the coefficients from the interaction between time and MNP exposure at baseline.

**Supplementary table S17.**
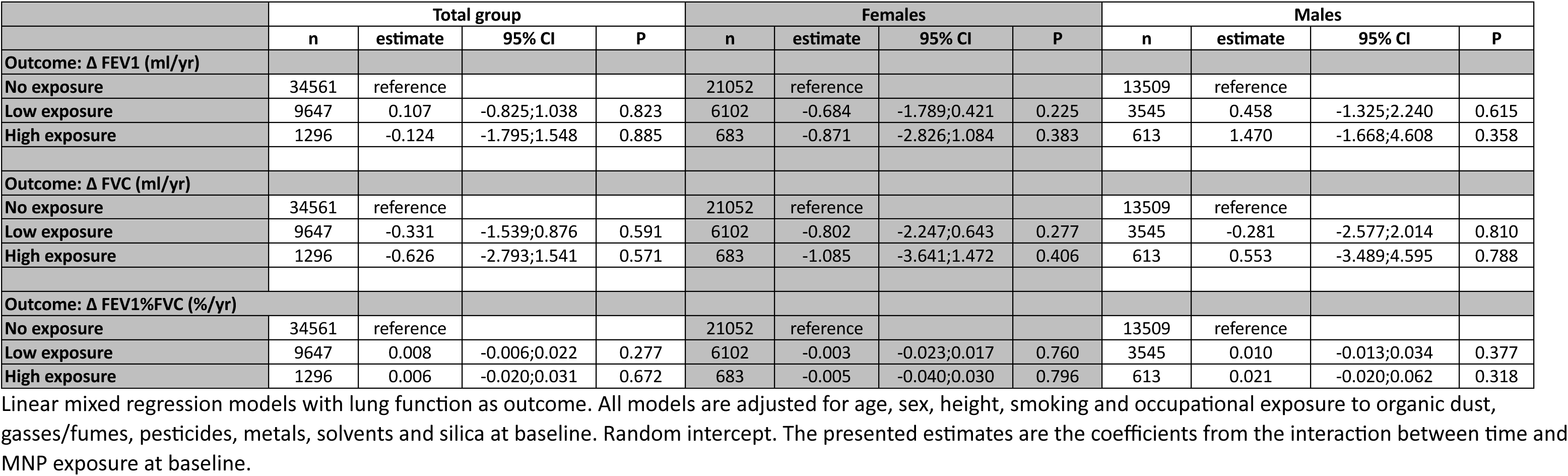
Associations between occupational MNP exposure and lung function decline – Main analysis - Total group and stratified by sex. Restricted to subjects with at least two lung function measurements and ≥ 25 years at baseline.

|  | Total group |  |  |  | Females |  |  |  | Males |  |  |  |
| --- | --- | --- | --- | --- | --- | --- | --- | --- | --- | --- | --- | --- |
|  | n | estimate | 95% CI | P | n | estimate | 95% CI | P | n | estimate | 95% CI | P |
| **Outcome: $\Delta$ FEV1 (ml/yr)** |  |  |  |  |  |  |  |  |  |  |  |  |
| No exposure | 34561 | reference |  |  | 21052 | reference |  |  | 13509 | reference |  |  |
| Low exposure | 9647 | 0.107 | -0.825;1.038 | 0.823 | 6102 | -0.684 | -1.789;0.421 | 0.225 | 3545 | 0.458 | -1.325;2.240 | 0.615 |
| High exposure | 1296 | -0.124 | -1.795;1.548 | 0.885 | 683 | -0.871 | -2.826;1.084 | 0.383 | 613 | 1.470 | -1.668;4.608 | 0.358 |
| **Outcome: $\Delta$ FVC (ml/yr)** |  |  |  |  |  |  |  |  |  |  |  |  |
| No exposure | 34561 | reference |  |  | 21052 | reference |  |  | 13509 | reference |  |  |
| Low exposure | 9647 | -0.331 | -1.539;0.876 | 0.591 | 6102 | -0.802 | -2.247;0.643 | 0.277 | 3545 | -0.281 | -2.577;2.014 | 0.810 |
| High exposure | 1296 | -0.626 | -2.793;1.541 | 0.571 | 683 | -1.085 | -3.641;1.472 | 0.406 | 613 | 0.553 | -3.489;4.595 | 0.788 |
| **Outcome: $\Delta$ FEV1%FVC (%/yr)** |  |  |  |  |  |  |  |  |  |  |  |  |
| No exposure | 34561 | reference |  |  | 21052 | reference |  |  | 13509 | reference |  |  |
| Low exposure | 9647 | 0.008 | -0.006;0.022 | 0.277 | 6102 | -0.003 | -0.023;0.017 | 0.760 | 3545 | 0.010 | -0.013;0.034 | 0.377 |
| High exposure | 1296 | 0.006 | -0.020;0.031 | 0.672 | 683 | -0.005 | -0.040;0.030 | 0.796 | 613 | 0.021 | -0.020;0.062 | 0.318 |

Linear mixed regression models with lung function as outcome. All models are adjusted for age, sex, height, smoking and occupational exposure to organic dust, gasses/fumes, pesticides, metals, solvents and silica at baseline. Random intercept. The presented estimates are the coefficients from the interaction between time and MNP exposure at baseline.

**Supplementary table S18.**
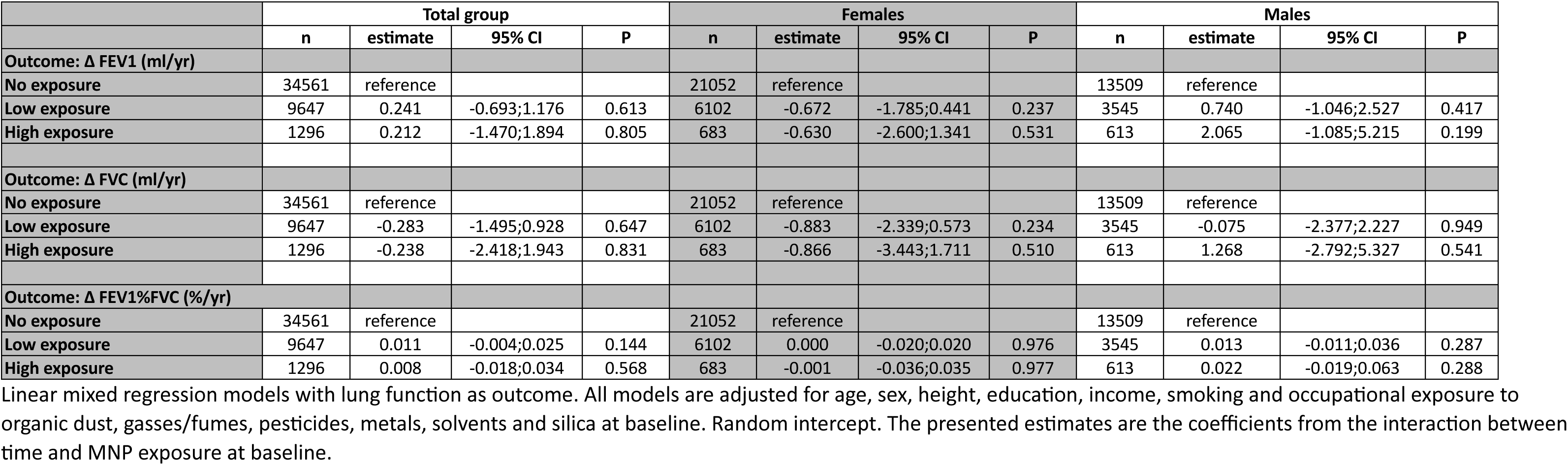
Associations between occupational MNP exposure and lung function decline – Sensitivity analysis: additional adjustment for education and income - Total group and stratified by sex. Restricted to subjects with at least two lung function measurements and ≥ 25 years at baseline.

**Supplementary table S19.** Associations between occupational MNP exposure and development of airway obstruction – Base analysis: no adjustment for co-exposures and no adjustment for education and income - Total group and stratified by sex.

|  | Total group |  |  |  | Females |  |  |  | Males |  |  |  |
| --- | --- | --- | --- | --- | --- | --- | --- | --- | --- | --- | --- | --- |
|  | n | OR | 95% CI | P | n | OR | 95% CI | P | n | OR | 95% CI | P |
| **Outcome: development of FEV1/FVC < 0.7** |  |  |  |  |  |  |  |  |  |  |  |  |
| No exposure | 31268 | reference |  |  | 19517 | reference |  |  | 11751 | reference |  |  |
| Low exposure | 8514 | 1.098 | 1.009;1.194 | 0.030 | 5505 | 1.107 | 0.994;1.232 | 0.064 | 3009 | 1.079 | 0.943;1.234 | 0.271 |
| High exposure | 1106 | 1.064 | 0.865;1.309 | 0.557 | 588 | 1.015 | 0.752;1.371 | 0.921 | 518 | 1.108 | 0.831;1.476 | 0.485 |
| **Outcome: development of FEV1/FVC < LLN** |  |  |  |  |  |  |  |  |  |  |  |  |
| No exposure | 32184 | reference |  |  | 19715 | reference |  |  | 12469 | reference |  |  |
| Low exposure | 8832 | 1.117 | 0.996;1.252 | 0.058 | 5644 | 1.197 | 1.041;1.376 | 0.012 | 3188 | 0.977 | 0.801;1.192 | 0.820 |
| High exposure | 1152 | 0.923 | 0.683;1.248 | 0.604 | 604 | 1.078 | 0.732;1.589 | 0.703 | 548 | 0.744 | 0.460;1.204 | 0.229 |

Logistic regression models with development of airway obstruction as outcome. All models are adjusted for age, sex, height, smoking and time between baseline and follow-up.

**Supplementary table S20.** Associations between occupational MNP exposure and development of airway obstruction – Main analysis - Total group and stratified by sex.

|  | Total group |  |  |  | Females |  |  |  | Males |  |  |  |
| --- | --- | --- | --- | --- | --- | --- | --- | --- | --- | --- | --- | --- |
|  | n | OR | 95% CI | P | n | OR | 95% CI | P | n | OR | 95% CI | P |
| **Outcome: development of FEV1/FVC < 0.7** |  |  |  |  |  |  |  |  |  |  |  |  |
| No exposure | 31268 | reference |  |  | 19517 | reference |  |  | 11751 | reference |  |  |
| Low exposure | 8514 | 1.026 | 0.905;1.163 | 0.686 | 5505 | 1.022 | 0.860;1.214 | 0.808 | 3009 | 1.009 | 0.825;1.234 | 0.931 |
| High exposure | 1106 | 1.032 | 0.824;1.294 | 0.782 | 588 | 0.981 | 0.717;1.342 | 0.905 | 518 | 1.130 | 0.799;1.599 | 0.490 |
| **Outcome: development of FEV1/FVC < LLN** |  |  |  |  |  |  |  |  |  |  |  |  |
| No exposure | 32184 | reference |  |  | 19715 | reference |  |  | 12469 | reference |  |  |
| Low exposure | 8832 | 0.979 | 0.823;1.164 | 0.808 | 5644 | 1.107 | 0.881;1.391 | 0.384 | 3188 | 0.818 | 0.610;1.097 | 0.180 |
| High exposure | 1152 | 0.842 | 0.608;1.166 | 0.842 | 604 | 1.039 | 0.691;1.562 | 0.856 | 548 | 0.680 | 0.386;1.201 | 0.184 |

Logistic regression models with development of airway obstruction as outcome. All models are adjusted for age, sex, height, smoking, time between baseline and follow-up and occupational exposure to organic dust, gasses/fumes, pesticides, metals, solvents and silica.

**Supplementary table S21.** Associations between occupational MNP exposure and development of airway obstruction – Sensitivity analysis: additional adjustment for education and income - Total group and stratified by sex.

|  | Total group |  |  |  | Females |  |  |  | Males |  |  |  |
| --- | --- | --- | --- | --- | --- | --- | --- | --- | --- | --- | --- | --- |
|  | n | OR | 95% CI | P | n | OR | 95% CI | P | n | OR | 95% CI | P |
| **Outcome: development of FEV1/FVC < 0.7** |  |  |  |  |  |  |  |  |  |  |  |  |
| No exposure | 31268 | reference |  |  | 19517 | reference |  |  | 11751 | reference |  |  |
| Low exposure | 8514 | 1.022 | 0.901;1.159 | 0.738 | 5505 | 1.023 | 0.860;1.218 | 0.796 | 3009 | 0.993 | 0.811;1.216 | 0.947 |
| High exposure | 1106 | 1.026 | 0.817;1.288 | 0.827 | 588 | 0.976 | 0.712;1.339 | 0.882 | 518 | 1.120 | 0.790;1.590 | 0.524 |
| **Outcome: development of FEV1/FVC < LLN** |  |  |  |  |  |  |  |  |  |  |  |  |
| No exposure | 32184 | reference |  |  | 19715 | reference |  |  | 12469 | reference |  |  |
| Low exposure | 8832 | 0.966 | 0.812;1.148 | 0.692 | 5644 | 1.105 | 0.878;1.391 | 0.397 | 3188 | 0.784 | 0.584;1.053 | 0.105 |
| High exposure | 1152 | 0.802 | 0.579;1.112 | 0.186 | 604 | 1.007 | 0.668;1.518 | 0.974 | 548 | 0.624 | 0.353;1.103 | 0.105 |

Logistic regression models with development of airway obstruction as outcome. All models are adjusted for age, sex, height, education, income, smoking, time between baseline and follow-up and occupational exposure to organic dust, gasses/fumes, pesticides, metals, solvents and silica.

**Supplementary table S21.** Associations between occupational MNP exposure and development of respiratory symptoms and asthma – Base analysis: no adjustment for co-exposures and no adjustment for education and income - Total group and stratified by sex.

|  | Total group |  |  |  | Females |  |  |  | Males |  |  |  |
| --- | --- | --- | --- | --- | --- | --- | --- | --- | --- | --- | --- | --- |
|  | n | OR | 95% CI | P | n | OR | 95% CI | P | n | OR | 95% CI | P |
| **Outcome: development of Wheeze** |  |  |  |  |  |  |  |  |  |  |  |  |
| No exposure | 47299 | reference |  |  | 29165 | reference |  |  | 18134 | reference |  |  |
| Low exposure | 13227 | 0.962 | 0.860;1.076 | 0.497 | 8590 | 0.962 | 0.833;1.110 | 0.593 | 4637 | 0.952 | 0.797;1.138 | 0.588 |
| High exposure | 1786 | 1.111 | 0.868;1.423 | 0.403 | 947 | 0.925 | 0.633;1.352 | 0.689 | 839 | 1.289 | 0.928;1.791 | 0.130 |
| **Outcome: development of Cough** |  |  |  |  |  |  |  |  |  |  |  |  |
| No exposure | 49716 | reference |  |  | 30618 | reference |  |  | 19098 | reference |  |  |
| Low exposure | 14031 | 1.040 | 0.964;1.122 | 0.311 | 9052 | 0.960 | 0.866;1.063 | 0.430 | 4979 | 1.158 | 1.033;1.298 | 0.012 |
| High exposure | 1883 | 1.210 | 1.022;1.433 | 0.027 | 1002 | 1.216 | 0.954;1.550 | 0.114 | 881 | 1.204 | 0.951;1.525 | 0.123 |
| **Outcome: development of Sputum** |  |  |  |  |  |  |  |  |  |  |  |  |
| No exposure | 50337 | reference |  |  | 31150 | reference |  |  | 19187 | reference |  |  |
| Low exposure | 14165 | 1.048 | 0.963;1.139 | 0.279 | 9198 | 0.964 | 0.859;1.081 | 0.526 | 4967 | 1.170 | 1.033;1.325 | 0.013 |
| High exposure | 1922 | 1.032 | 0.847;1.258 | 0.752 | 1019 | 0.871 | 0.637;1.190 | 0.385 | 903 | 1.184 | 0.916;1.530 | 0.196 |
| **Outcome: development of Bronchitis** |  |  |  |  |  |  |  |  |  |  |  |  |
| No exposure | 52008 | reference |  |  | 32021 | reference |  |  | 19987 | reference |  |  |
| Low exposure | 14673 | 1.127 | 1.022;1.243 | 0.016 | 9467 | 1.038 | 0.909;1.185 | 0.582 | 5206 | 1.261 | 1.089;1.459 | 0.002 |
| High exposure | 2007 | 1.066 | 0.847;1.343 | 0.586 | 1053 | 0.818 | 0.561;1.193 | 0.298 | 954 | 1.301 | 0.969;1.745 | 0.080 |
| **Outcome: development of Asthma** |  |  |  |  |  |  |  |  |  |  |  |  |
| No exposure | 49343 | reference |  |  | 30286 | reference |  |  | 19057 | reference |  |  |
| Low exposure | 13870 | 1.089 | 0.990;1.198 | 0.081 | 8888 | 1.062 | 0.942;1.197 | 0.325 | 4982 | 1.172 | 1.000;1.374 | 0.050 |
| High exposure | 1905 | 1.141 | 0.914;1.426 | 0.244 | 998 | 1.298 | 0.978;1.722 | 0.071 | 907 | 0.986 | 0.686;1.418 | 0.940 |

Logistic regression models with development of symptoms and asthma as outcome. All models are adjusted for age, sex, BMI and smoking.

**Supplementary table S22.** Associations between occupational MNP exposure and development of respiratory symptoms and asthma – Main analysis - Total group and stratified by sex.

|  | Total group |  |  |  | Females |  |  |  | Males |  |  |  |
| --- | --- | --- | --- | --- | --- | --- | --- | --- | --- | --- | --- | --- |
|  | n | OR | 95% CI | P | n | OR | 95% CI | P | n | OR | 95% CI | P |
| **Outcome: development of Wheeze** |  |  |  |  |  |  |  |  |  |  |  |  |
| No exposure | 47299 | reference |  |  | 29165 | reference |  |  | 18134 | reference |  |  |
| Low exposure | 13227 | 0.953 | 0.812;1.118 | 0.556 | 8590 | 0.881 | 0.706;1.100 | 0.263 | 4637 | 1.086 | 0.843;1.399 | 0.523 |
| High exposure | 1786 | 1.132 | 0.866;1.479 | 0.365 | 947 | 0.893 | 0.602;1.324 | 0.573 | 839 | 1.578 | 1.063;2.343 | 0.024 |
| **Outcome: development of Cough** |  |  |  |  |  |  |  |  |  |  |  |  |
| No exposure | 49716 | reference |  |  | 30618 | reference |  |  | 19098 | reference |  |  |
| Low exposure | 14031 | 0.965 | 0.863;1.079 | 0.537 | 9052 | 0.943 | 0.802;1.108 | 0.473 | 4979 | 1.033 | 0.872;1.224 | 0.706 |
| High exposure | 1883 | 1.132 | 0.941;1.362 | 0.188 | 1002 | 1.209 | 0.939;1.557 | 0.142 | 881 | 1.083 | 0.816;1.439 | 0.580 |
| **Outcome: development of Sputum** |  |  |  |  |  |  |  |  |  |  |  |  |
| No exposure | 50337 | reference |  |  | 31150 | reference |  |  | 19187 | reference |  |  |
| Low exposure | 14165 | 0.970 | 0.858;1.096 | 0.622 | 9198 | 0.894 | 0.749;1.068 | 0.217 | 4967 | 1.138 | 0.948;1.367 | 0.166 |
| High exposure | 1922 | 0.976 | 0.786;1.211 | 0.824 | 1019 | 0.839 | 0.606;1.160 | 0.288 | 903 | 1.131 | 0.829;1.544 | 0.437 |
| **Outcome: development of Bronchitis** |  |  |  |  |  |  |  |  |  |  |  |  |
| No exposure | 52008 | reference |  |  | 32021 | reference |  |  | 19987 | reference |  |  |
| Low exposure | 14673 | 0.976 | 0.846;1.126 | 0.736 | 9467 | 0.893 | 0.725;1.099 | 0.285 | 5206 | 1.162 | 0.936;1.441 | 0.173 |
| High exposure | 2007 | 0.948 | 0.736;1.221 | 0.679 | 1053 | 0.764 | 0.517;1.129 | 0.177 | 954 | 1.160 | 0.810;1.660 | 0.419 |
| **Outcome: development of Asthma** |  |  |  |  |  |  |  |  |  |  |  |  |
| No exposure | 49343 | reference |  |  | 30286 | reference |  |  | 19057 | reference |  |  |
| Low exposure | 13870 | 1.020 | 0.886;1.174 | 0.783 | 8888 | 0.999 | 0.830;1.203 | 0.994 | 4982 | 1.077 | 0.850;1.366 | 0.538 |
| High exposure | 1905 | 1.142 | 0.901;1.448 | 0.273 | 998 | 1.284 | 0.957;1.722 | 0.096 | 907 | 0.948 | 0.620;1.452 | 0.807 |

Logistic regression models with development of symptoms and asthma as outcome. All models are adjusted for age, sex, BMI, smoking and occupational exposure to organic dust, gasses/fumes, pesticides, metals, solvents and silica.

**Supplementary table S23.** Associations between occupational MNP exposure and development of respiratory symptoms and asthma – Sensitivity analysis: additional adjustment for education and income - Total group and stratified by sex.

|  | Total group |  |  |  | Females |  |  |  | Males |  |  |  |
| --- | --- | --- | --- | --- | --- | --- | --- | --- | --- | --- | --- | --- |
|  | n | OR | 95% CI | P | n | OR | 95% CI | P | n | OR | 95% CI | P |
| **Outcome: development of Wheeze** |  |  |  |  |  |  |  |  |  |  |  |  |
| No exposure | 47299 | reference |  |  | 29165 | reference |  |  | 18134 | reference |  |  |
| Low exposure | 13227 | 0.925 | 0.789;1.085 | 0.336 | 8590 | 0.848 | 0.679;1.060 | 0.147 | 4637 | 1.030 | 0.800;1.327 | 0.819 |
| High exposure | 1786 | 1.028 | 0.786;1.344 | 0.843 | 947 | 0.809 | 0.545;1.201 | 0.292 | 839 | 1.424 | 0.958;2.115 | 0.080 |
| **Outcome: development of Cough** |  |  |  |  |  |  |  |  |  |  |  |  |
| No exposure | 49716 | reference |  |  | 30618 | reference |  |  | 19098 | reference |  |  |
| Low exposure | 14031 | 0.939 | 0.840;1.050 | 0.272 | 9052 | 0.908 | 0.772;1.068 | 0.243 | 4979 | 0.995 | 0.840;1.179 | 0.952 |
| High exposure | 1883 | 1.043 | 0.866;1.256 | 0.656 | 1002 | 1.094 | 0.848;1.410 | 0.490 | 881 | 1.019 | 0.766;1.354 | 0.899 |
| **Outcome: development of Sputum** |  |  |  |  |  |  |  |  |  |  |  |  |
| No exposure | 50337 | reference |  |  | 31150 | reference |  |  | 19187 | reference |  |  |
| Low exposure | 14165 | 0.939 | 0.831;1.061 | 0.313 | 9198 | 0.851 | 0.712;1.017 | 0.075 | 4967 | 1.101 | 0.916;1.322 | 0.305 |
| High exposure | 1922 | 0.891 | 0.718;1.107 | 0.298 | 1019 | 0.747 | 0.540;1.035 | 0.079 | 903 | 1.063 | 0.778;1.451 | 0.703 |
| **Outcome: development of Bronchitis** |  |  |  |  |  |  |  |  |  |  |  |  |
| No exposure | 52008 | reference |  |  | 32021 | reference |  |  | 19987 | reference |  |  |
| Low exposure | 14673 | 0.951 | 0.824;1.096 | 0.487 | 9467 | 0.862 | 0.699;1.062 | 0.162 | 5206 | 1.127 | 0.909;1.398 | 0.276 |
| High exposure | 2007 | 0.883 | 0.686;1.138 | 0.337 | 1053 | 0.703 | 0.476;1.040 | 0.078 | 954 | 1.106 | 0.772;1.583 | 0.584 |
| **Outcome: development of Asthma** |  |  |  |  |  |  |  |  |  |  |  |  |
| No exposure | 49343 | Reference |  |  | 30286 | reference |  |  | 19057 | reference |  |  |
| Low exposure | 13870 | 1.002 | 0.871;1.153 | 0.979 | 8888 | 0.968 | 0.803;1.165 | 0.729 | 4982 | 1.049 | 0.828;1.329 | 0.692 |
| High exposure | 1905 | 1.066 | 0.840;1.353 | 0.598 | 998 | 1.172 | 0.872;1.574 | 0.294 | 907 | 0.898 | 0.587;1.374 | 0.620 |

Logistic regression models with development of symptoms and asthma as outcome. All models are adjusted for age, sex, BMI, education, income, smoking and occupational exposure to organic dust, gasses/fumes, pesticides, metals, solvents and silica.

## Supplemental figures

**Supplementary figure S1:**
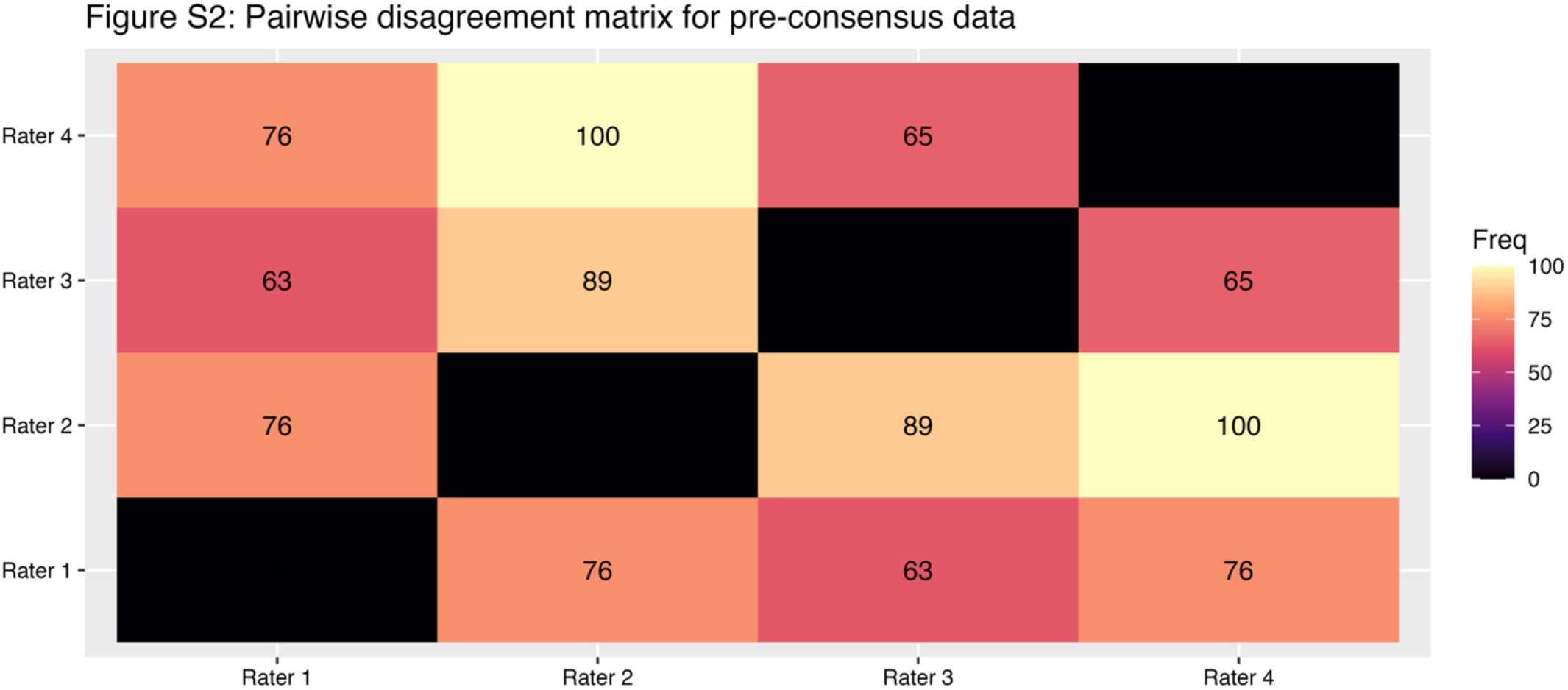
Pairwise disagreement matrix of pre-consensus data. Total number of jobs rated differently between the four raters in pairwise combinations.

**Supplementary figure S2:**
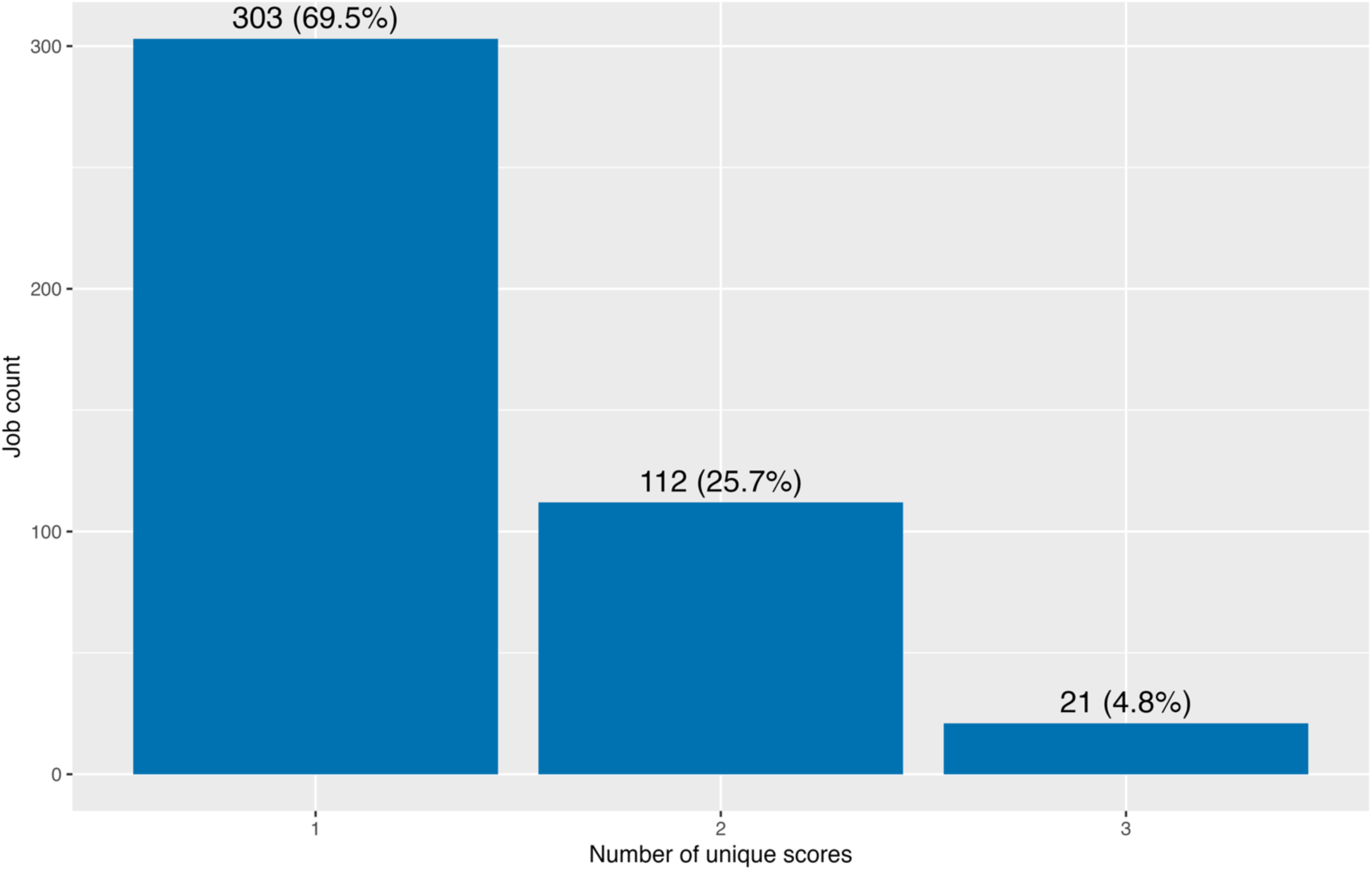
DistribuGon of jobs by agreement level. Total number of jobs that had 1, 2, or 3 unique scores for the four different raters. Numbers on top of the bars indicate total number of jobs (percentage of all jobs).

**Supplementary figure S3:**
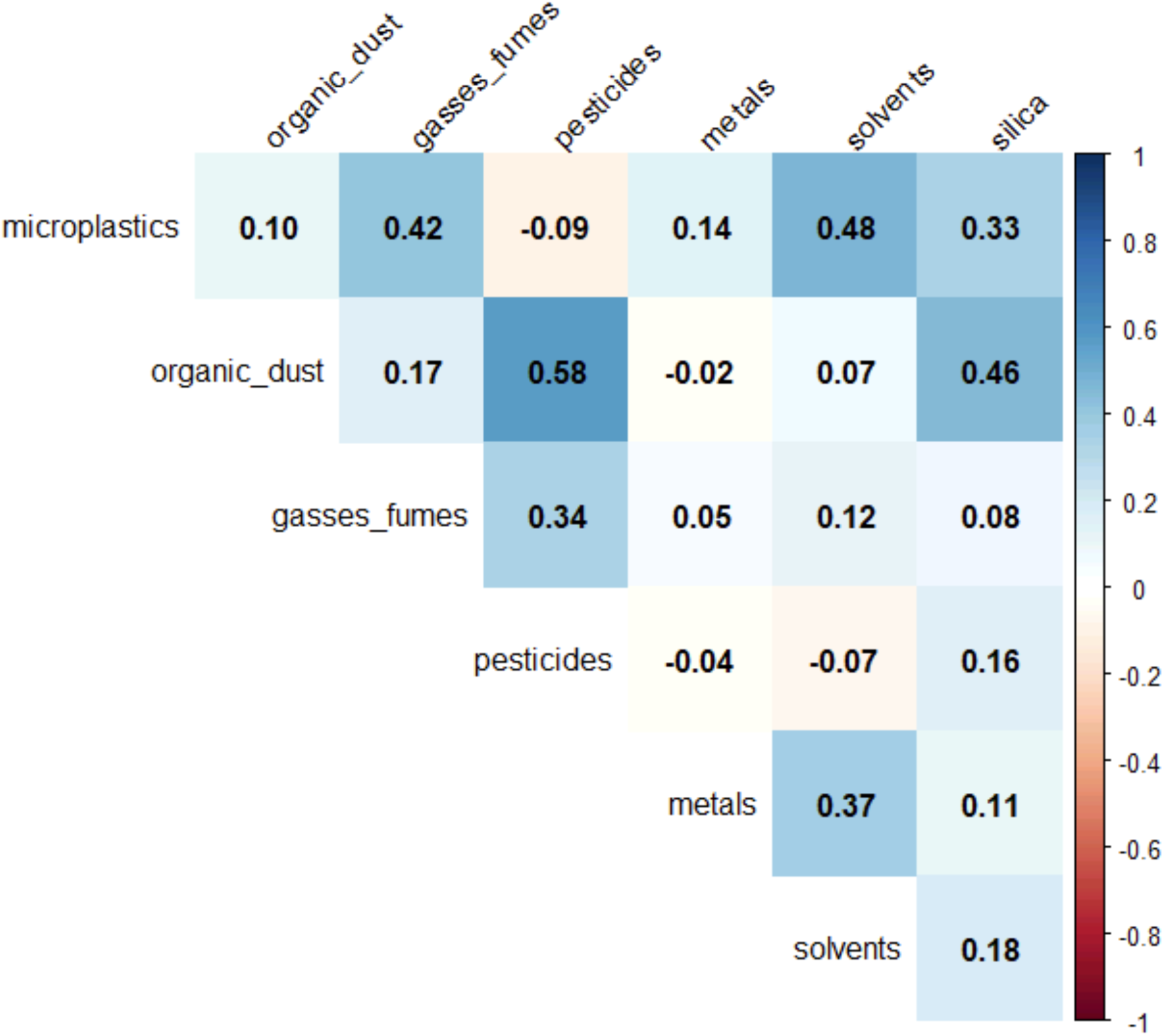
Spearman rank-correlaGons between occupaGonal MNP exposure and the other occupaGonal exposures in all subjects with exposure data available (n=136,928).

**Supplementary figure S4:**
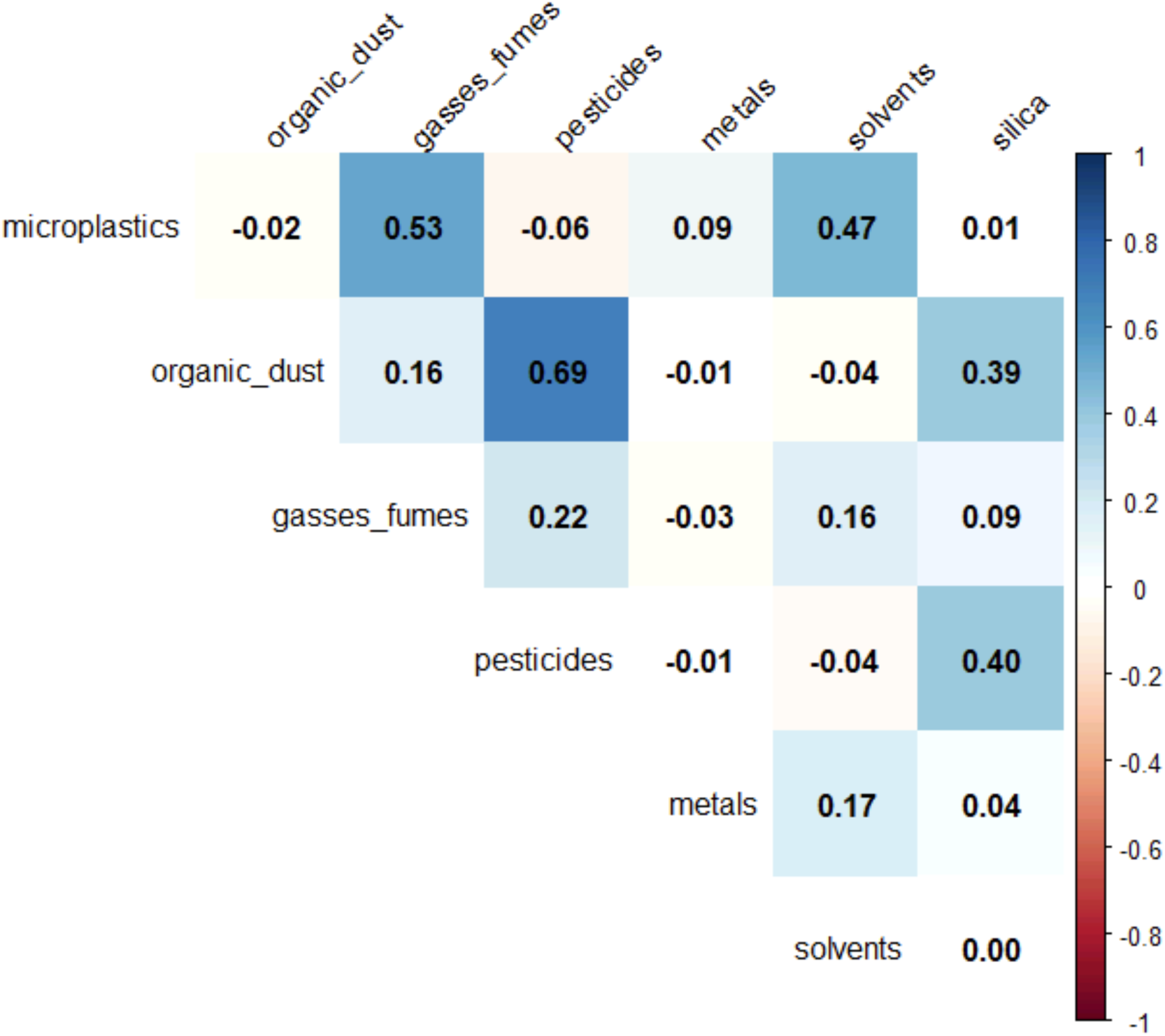
Spearman rank-correlaGons between occupaGonal MNP exposure and the other occupaGonal exposures in all female subjects with exposure data available (n=83,669).

**Supplementary figure S5:**
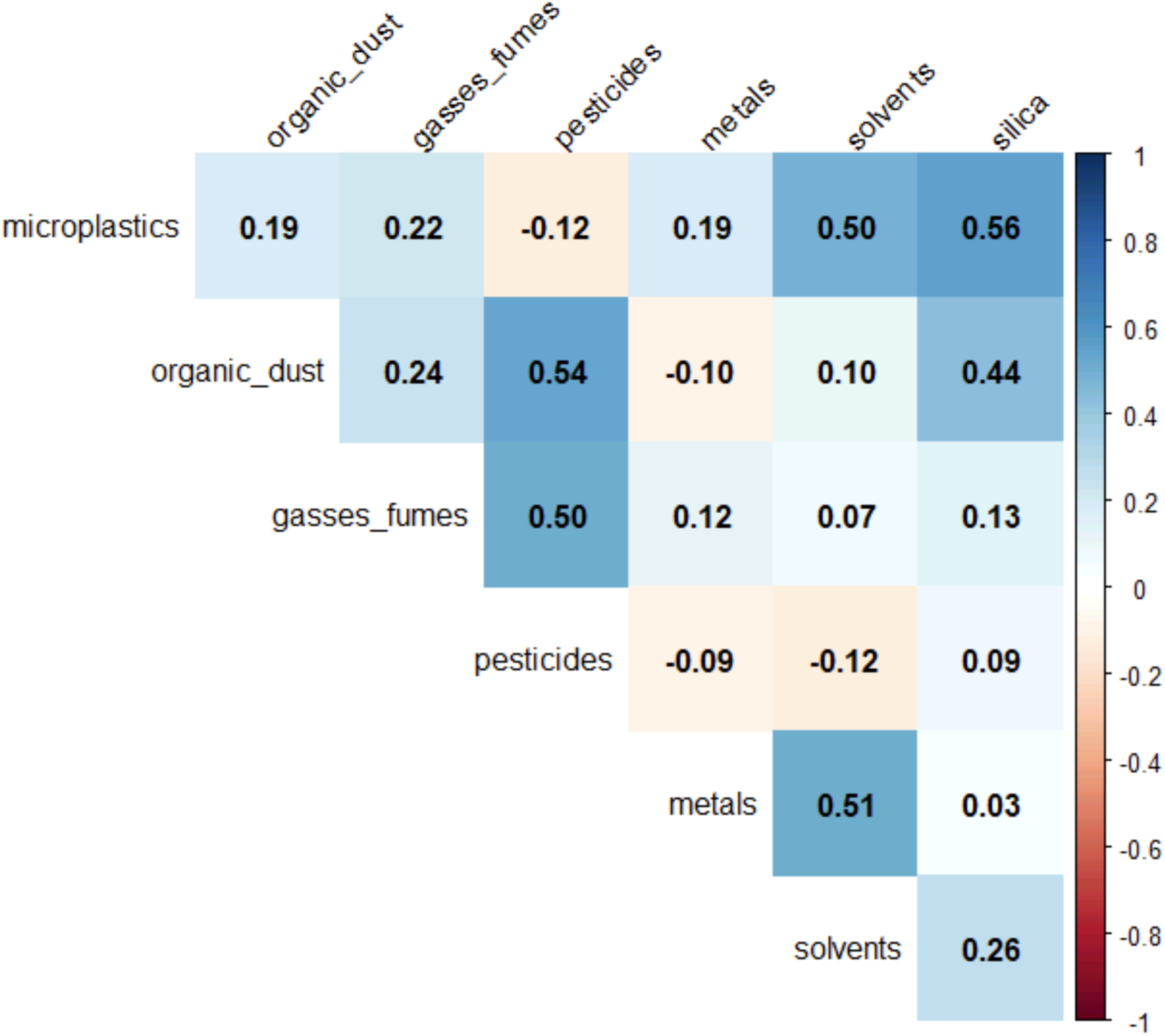
Spearman rank-correlaGons between occupaGonal MNP exposure and the other occupaGonal exposures in all male subjects with exposure data available (n=53,256).

## Notes

### Competing Interest Statement

Barbro Melgert, Gwenda Vasse, Nienke Vrisekoop, and Joelle Klazen report financial support was provided by Netherlands Organisation for Health Research and Development. Barbro Melgert has intellectual property #PlastiXJEM (job-exposure matrix for occupational airborne micro-and nanoplastics) deposited in the name of Barbro Melgert (BOIP i-DEPOT No. 157917). All authors declare that they have no known competing financial interests or personal relationships that could have appeared to influence the work reported in this manuscript.

### Author Declarations

The Medical Ethics Committee (METc) of the University Medical Center Groningen, The Netherlands approved the study under approval number METC 2007/152. All participants provided written informed consent.

### Summary of Updates

The introduction was streamlined and now ends with a concise aim statement. The Methods section clarifies the exposure assignment approach, adds a definition of discrepancies in the consensus process, and moves the correlation method description from Results. Longitudinal analyses were extended with lung function data from the third Lifelines health assessment, increasing the sample size from 24,106 to 45,504 participants using linear mixed effects models. The Results section now explicitly highlights dose-dependent patterns, includes sector-specific sensitivity analyses, adds sex-stratified results to Figure 5, and presents longitudinal results in a new Table 2. The Discussion now opens with the absence of longitudinal associations, includes a new paragraph on clinical relevance of effect sizes, and adds the healthy worker effect and exposure changes during follow-up as limitations. The abstract was revised to include the dose-dependent pattern and remove the sensitivity analysis. The Lifelines cohort description and funding acknowledgment were added per Lifelines requirements.

